# Reverse engineering of motor unit discharge in multiple sclerosis reveals heterogeneity of voluntary motor commands

**DOI:** 10.64898/2026.06.15.26355613

**Authors:** Laura M. McPherson, Keith R. Lohse, Skyler M. Simon, Daniel B. Free, James A. Beauchamp, Francesco Negro, Robert T. Naismith, Anne H. Cross

**Affiliations:** Program in Physical Therapy, Washington University School of Medicine, St. Louis, MO; Department of Neurology, Washington University School of Medicine, St. Louis, MO; Department of Mechanical Engineering, Carnegie Mellon University, Pittsburgh, PA; Department of Clinical and Experimental Sciences, Universita degli Studi di Brescia, Brescia, IT

**Keywords:** Multiple sclerosis, motor unit, motor control, neuroscience, neurophysiology, motoneuron, rehabilitation

## Abstract

Central nervous system injury causes motor deficits through derangement of excitatory, inhibitory, and/or neuromodulatory inputs to motoneurons, the three fundamental components of motor commands. Typically, study of pathologic neural control in humans is restricted to only one of the three. Chardon et al. (2024) presented a fundamentally new approach to comprehensively study all components by reverse engineering motor unit firing patterns. We apply their framework to motor unit firing patterns from 89 people with multiple sclerosis (MS) and 34 controls to study excitatory, inhibitory, and neuromodulatory contributions to pathologic motor output. Disruptions to all components are plausible in MS, a disease hallmarked by heterogeneity in nearly all aspects. Accordingly, we found abnormalities in MS for all three components. Notably, neuromodulation included both high and low extremes. Our results suggest that pathophysiology of motor commands in MS varies among patients, a finding fundamentally different from other studied populations showing relative consistency.

## 2. Introduction

Central nervous system (CNS) injury causes motor deficits through the derangement of excitatory, inhibitory, and/or neuromodulatory inputs to spinal motoneurons. These inputs converge on the motoneuron from all areas of the neuraxis and make up the three fundamental components of voluntary motor commands that are necessary for skilled motor control. Excitatory and inhibitory inputs from cortical, afferent, and spinal interneuronal sources convey specific task-related commands to motoneurons (Heckman & Enoka, 2012).

Neuromodulatory inputs, delivered as monoamines from descending brainstem pathways, dramatically transform motoneuron excitability to meet task demands, amplifying a motoneuron’s response to excitatory and inhibitory inputs (Heckman *et al*., 2009; Heckman & Enoka, 2012). The synaptic organization of the three inputs dictates the specific pattern of motoneuron firing that results and is thus highly relevant to motor behavior.

Recent technical and computational advances have transformed our ability to estimate components of the motor command in humans through recordings from motor units, whose discharge has a 1:1 relationship with that of their motoneurons. High-density EMG arrays and decomposition algorithms deliver detailed spiking patterns of *populations* of simultaneously firing motoneurons in humans, offering enormous potential for uncovering deep insights into the organization of motor commands. Chardon et al. recently developed the first approach to take advantage of this potential, capitalizing on realistic motoneuron models established from extensive study of motoneuron input-output functions in animal preparations (Chardon *et al*., 2024). Using seven temporal and geometric features extracted from simulated motor unit firing patterns with known excitatory, inhibitory, and neuromodulatory inputs, their reverse engineering model successfully estimated the simulated patterns of inputs with a high level of accuracy. Importantly, their simulations showed that no one firing pattern feature was sufficient for estimating the organization of synaptic input.

Despite these advances, the study of motor control following CNS injury in humans is typically restricted to only one of the three motor command components. Here, we apply the reverse engineering framework to a robust sample of motor unit firing patterns from people with multiple sclerosis (MS) as a means to study excitatory, inhibitory, and neuromodulatory contributions to pathologic motor output.

MS is an immune-mediated inflammatory disease characterized by demyelination and neurodegeneration in the brain and spinal cord (Compston & Coles, 2008; Oh, 2022). Because MS causes CNS damage that is multi-focal and scattered across many structures, disruptions to all three components of the motor command are plausible. Additionally, MS is hallmarked by wide variation from person to person in nearly all aspects of the disease, including the areas of the CNS affected, the characteristics of lesions, and the sensorimotor impairments that result (Disanto *et al*., 2011; Oh, 2022). Thus, the potential for high variability across individuals in how motor commands are affected in MS necessitates a comprehensive approach like the reverse engineering framework of Chardon et. al (2024).

Our overall hypothesis was that voluntary motor commands are disrupted in different ways for different patients with MS. To test this hypothesis, we sought to understand how excitatory, inhibitory, and neuromodulatory components of the motor command, estimated using the reverse engineering framework, were distributed in our participants with MS relative to those from neurologically intact controls. If voluntary motor commands in the MS group are disrupted in the same way among patients, we reasoned that the values from the MS group would all be shifted in the same direction compared to controls. However, if they are disrupted in different ways among patients, as we hypothesized, we reasoned that MS distributions could exhibit values at both high and low extremes and may have multiple peaks in the distributions.

We applied the reverse engineering framework from Chardon et al. (2024) to motor unit firing patterns of the tibialis anterior and soleus muscles of 89 participants with MS who demonstrated a range of sensorimotor symptoms and disability and 34 neurologically intact controls. In support of our hypothesis, our results suggest that, among individuals with MS, excitatory, inhibitory, and neuromodulatory components of the motor command are disrupted in a variety of ways, with only a few areas of consistency. This finding is consistent with the heterogeneity of lesion locations and clinical symptoms observed in MS and provides a solid basis for further investigations into the nature of disrupted motor commands experienced by individuals—or perhaps subgroups of individuals—with MS. Importantly, our study demonstrates that the reverse engineering framework of Chardon et al. can be feasibly and meaningfully applied to human data to reveal new insights that were otherwise inaccessible.

## 3. Results

### 3.1. Application of Reverse Engineering Approach in Humans

To generate motor unit firing patterns suitable for reverse engineering, participants performed a slow “triangle” muscle contraction to parallel the simulated excitatory triangle trajectory (Johnson *et al*., 2017; Chardon *et al*., 2024). We tested 89 participants with MS (with disability ranging from none to severe) and 34 controls without neurological injury (matched by sex and age within 5 years) (see *Methods* for summary of demographics).

Participants performed the triangle contraction while sitting in a Biodex experimental chair with the tested ankle affixed to the accompanying dynamometer (**Figure 1**). First, their maximum voluntary torque (MVT) was measured in dorsiflexion and plantarflexion (results presented in **Appendix 1 - Figure A1**). Then, guided by real-time visual feedback of torque performance, they completed the triangle contraction by increasing their isometric dorsiflexion or plantarflexion torque to 20% of their MVT over 10 seconds and then decreasing the torque back to rest over another 10 seconds. High-density surface EMG was recorded from the tibialis anterior and soleus muscles. Offline, EMG was decomposed into individual motor unit spike trains using blind source separation (Negro *et al*., 2016), which were then smoothed using support vector regression to obtain a continuous firing pattern profile (Beauchamp *et al*., 2022).

**Figure 1.**
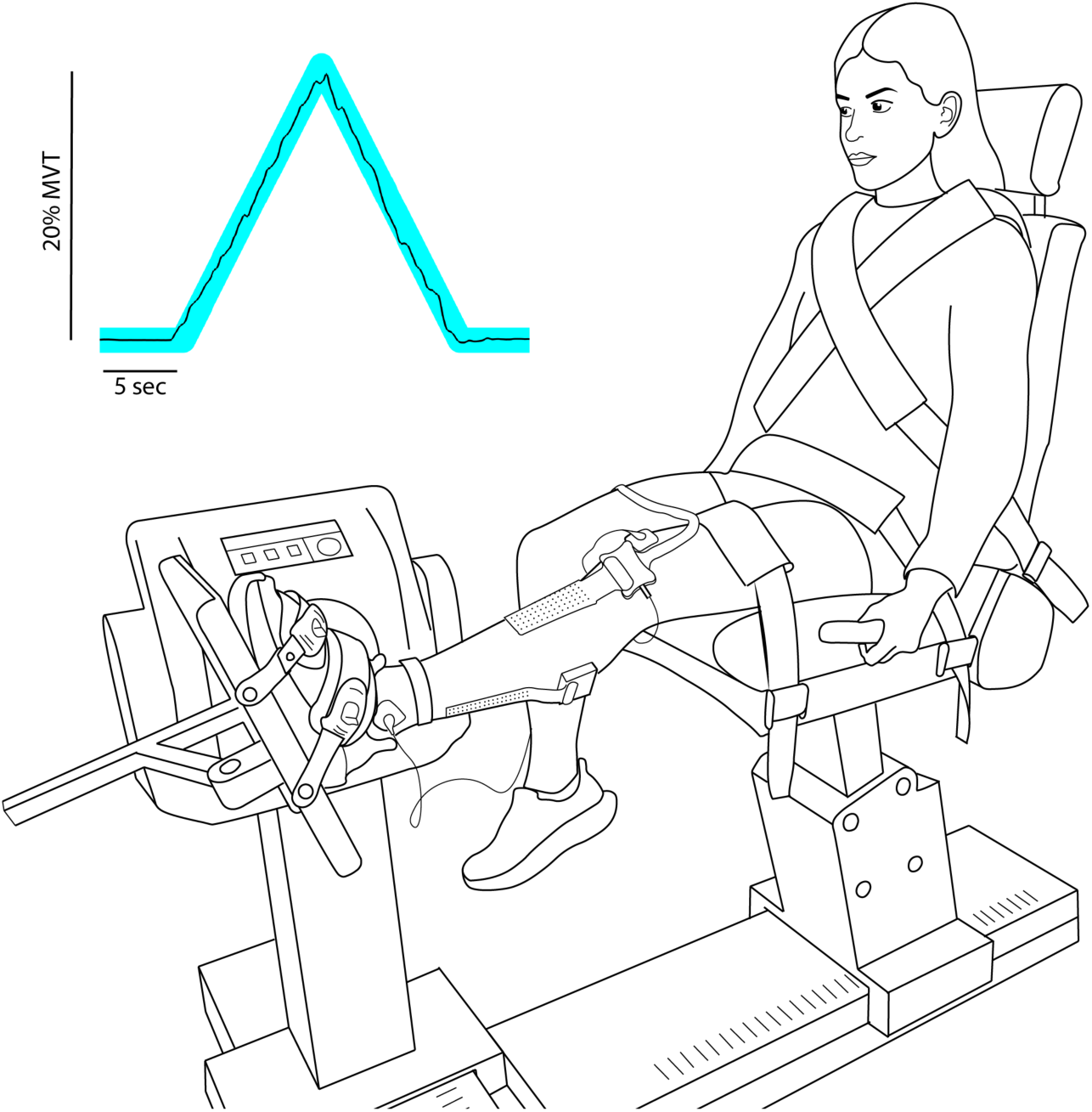
Experimental setup. Participants were seated with the tested foot secured to the dynamometer. High-density surface EMG arrays were affixed to the TA and soleus muscles. Real-time visual feedback of torque generation during experimental trials (inset) was displayed on a TV monitor.

For each continuous motor unit firing pattern, we extracted the seven features used by Chardon et al (**Figure 2A and 2B**). Several of the features are calculated relative to the time course of the simulated excitatory triangle trajectory (recruitment time, de-recruitment time, rate attenuation slope, braceheight). Because our data is from humans rather than simulated, we calculated these features relative to the participant’s torque trajectory rather than time. This modification captures the participant’s actual trajectory, given that using time would represent a perfectly performed triangle contraction. Calculation of these features is shown in **Figures 2A** and **2B** and detailed in *Methods*. In addition, we used the normalized version of delta-F to control for the effects of firing rate modulation on unnormalized delta-F values (Škarabot *et al*., 2025).

**Figure 2.**
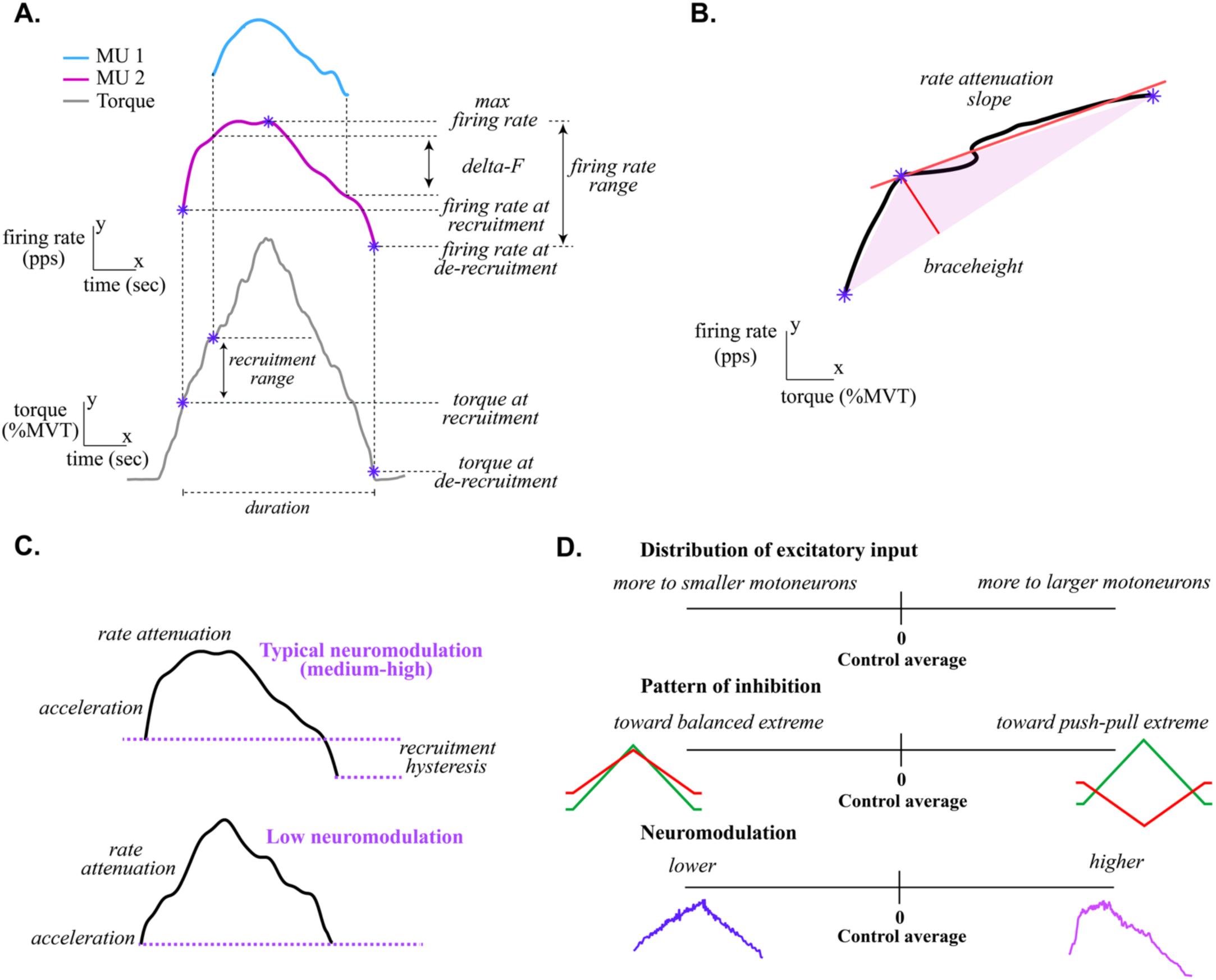
Calculation of reverse engineering features and interpretation of composite variables. A. Calculation of reverse engineering features from the smoothing firing rate vs. time function: torque at recruitment and de-recruitment, duration, recruitment range, delta-F, maximal firing rate, firing rate at recruitment and de-recruitment, and firing rate range. The delta-F calculation shown is for the higher threshold blue motor unit, whereas the other features are shown for the lower threshold purple motor unit. **B. Calculation of reverse engineering features from the smoothed firing rate vs. torque function:** braceheight and rate attenuation slope. **C. Neuromodulatory input facilitates persistent inward currents PICs) that drastically affect motor unit firing patterns.** Top: during a triangular contraction, motor units with large PICs produce firing patterns with a characteristic non-linear shape. Three distinct phases can be seen: rapid initial acceleration, rate attenuation marked by a more gradual rate increase, and recruitment hysteresis where the motor unit de-recruits at a lower level of excitatory synaptic input than when it was recruited. The example shown is a motor unit from the TA of a control participant. Bottom: Motor units with small (or no) PICs produce more linear firing patterns. **D. Physiologic interpretation of excitatory, inhibitory, and neuromodulatory composite variables.**

The final seven features were: firing rate non-linearity during ascent (*braceheight*), firing rate hysteresis (*normalized delta-F*), firing rate saturation (*rate attenuation slope*), torque at recruitment, torque at de-recruitment, recruitment range, and firing duration. We also report values of unnormalized delta-F for completeness.

Then, for each participant, we calculated a composite score for each type of synaptic input (excitation, inhibition, neuromodulation). First, we calculated one aggregate value for each reverse engineering feature per participant (see *Methods*) and converted it to a Z-score relative to the control group mean and standard deviation. Then, the composite score for each component was calculated using a weighted average of the reverse engineering features contributing to prediction of that input (**Table 1**). The weights for each feature were based on the features’ mutual information score with a given component, as determined by Chardon et al. (2024), with signs added to denote the relationships between variables (Powers *et al*., 2012; Beauchamp *et al*., 2023; Chardon *et al*., 2024). Details of the composite score calculation are outlined in *Methods*. We also calculated four key firing rate characteristics (maximum firing rate, firing rate range, firing rate at recruitment, and firing rate at de-recruitment) which we averaged to create a firing rate composite score.

**Table 1.** Weights for each reverse engineering feature in calculating excitation, inhibition, and neuromodulation composite variables.

|  | Excitation | Inhibition | Neuromodulation |
| --- | --- | --- | --- |
| Braceheight | -0.30 |  | 0.76 |
| Delta-F | -0.02 | 0.16 | 0.60 |
| Rate attenuation slope | 0.20 | 0.13 | -0.14 |
| Torque at recruitment | -0.43 |  | 0.30 |
| Torque at de-recruitment | -0.93 |  | -0.24 |
| Duration | 0.63 |  | 0.09 |
| Recruitment Range | -0.81 | 0.03 |  |

### 3.2. Characteristics of excitation, inhibition, and neuromodulation estimated by reverse engineering

The reverse engineering model estimates different characteristics of the three motor command components: (1) the *relative distribution of excitatory synaptic input* across motoneurons of different size (e.g., slow, smaller, lower-threshold vs. fast, larger, higher-threshold), (2) the *level of neuromodulatory input* (e.g., low, high), and (3) the *pattern of inhibitory input* relative to excitatory input over time (e.g., proportional vs. reciprocal).

Our composite variable for excitation quantifies the time course of sequential motoneuron recruitment, giving information about the relative amount of excitatory current to smaller (slow) vs. larger (fast) motoneurons delivered by various sources of excitatory input. Chardon et al. (2024) referred to this as the excitatory weight ratio. Motoneurons are recruited in order of increasing size (i.e., Henneman’s size principle), and if they received an equal amount of excitatory current, the spacing between sequential recruitment would be governed only by the motoneurons’ intrinsic electrical properties. However, excitatory inputs do not project uniformly to motoneurons of all sizes, so they lower the recruitment thresholds of the motoneurons to which they deliver more current (Powers & Binder, 2001; Binder *et al*., 2002; Johnson *et al*., 2017; Chardon *et al*., 2024). A value of zero on our excitation composite variable corresponds to the control group average. A positive value indicates relatively more input to larger motoneurons compared with the control group average, and a negative value indicates relatively more input to smaller motoneurons compared with the control group average.

The neuromodulation composite variable reflects the level of neuromodulatory input to motoneurons from descending monoaminergic brainstem pathways. Neuromodulatory input profoundly increases motoneuron excitability through facilitation of dendritic persistent inward currents (PICs) that change how the motoneuron processes and responds to excitatory and inhibitory inputs (i.e., its input/output function) (Heckman *et al*., 2005, 2009; Heckman & Enoka, 2012). PICs result in motoneuron firing patterns that are highly non-linear in relation to a linear excitatory input, with a rapid initial acceleration of firing rate, followed by an attenuation of firing rate increase, and then recruitment hysteresis (**Figure 2C**). A value of zero for our neuromodulation composite variable corresponds to the control group average, a positive value indicates relatively more neuromodulatory input, and a negative value indicates relatively less neuromodulatory input.

The inhibition composite variable gives information about the pattern of inhibitory input relative to the increasing and decreasing excitatory input. Chardon et al. (2024) modeled inhibition relative to excitation as a spectrum, ranging from changing proportionally (a “proportional” or “balanced” excitation-inhibition scheme) to changing reciprocally (a “push-pull” excitation-inhibition scheme) (**Figure 2D**). These two patterns of inhibition have strong effects on the linearity of motor unit firing patterns because of the potent effect of inhibition on PICs (Powers *et al*., 2012). The push-pull scheme, in which inhibition *decreases* from a tonic background level as excitation increases, allows stronger PIC amplitudes and results in more non-linear firing patterns. The balanced scheme, in which inhibition *increases* as excitation increases, limits PIC amplitude and reduces the net excitatory synaptic drive, resulting in sharply reduced rate modulation. A value of zero for our inhibition composite variable corresponds to the control group average, which is likely at some level of push-pull inhibition for the TA (Chardon *et al*., 2024). A positive value for the inhibition composite indicates inhibition that is closer to the push-pull scheme end of the spectrum, and a negative value indicates inhibition that is closer to the balanced end of the spectrum.

### 3.3. Task feasibility and individual motor unit firing profiles

Participants in the MS group were generally able to perform the triangle task with sufficient smoothness and symmetry, with the exception of three participants who were not able to complete the dorsiflexion task because of profound weakness and little-to-no voluntary control of their dorsiflexors (soleus data is included for these participants). Otherwise, compared with control participants, it sometimes required more trials for MS participants to achieve acceptable torque profiles.

There was a wide range of motor unit firing profiles in the MS group, and the variability can be noted visually by the three examples in **Figure 3**. All three MS participants whose data is shown had both supraspinal and spinal lesions. Motor unit firing profiles from the control participant (age between 26-30 yo) are typical for a young adult for both muscles (Beauchamp *et al*., 2022). S01 was of a similar age (between 31-35 yo) and did not yet have motor deficits, but the firing profiles in their TA motor units are noticeably different: a compressed recruitment range, significant self-sustained firing after cessation of the task, and notable synchrony among motor unit firing patterns.

**Figure 3.**
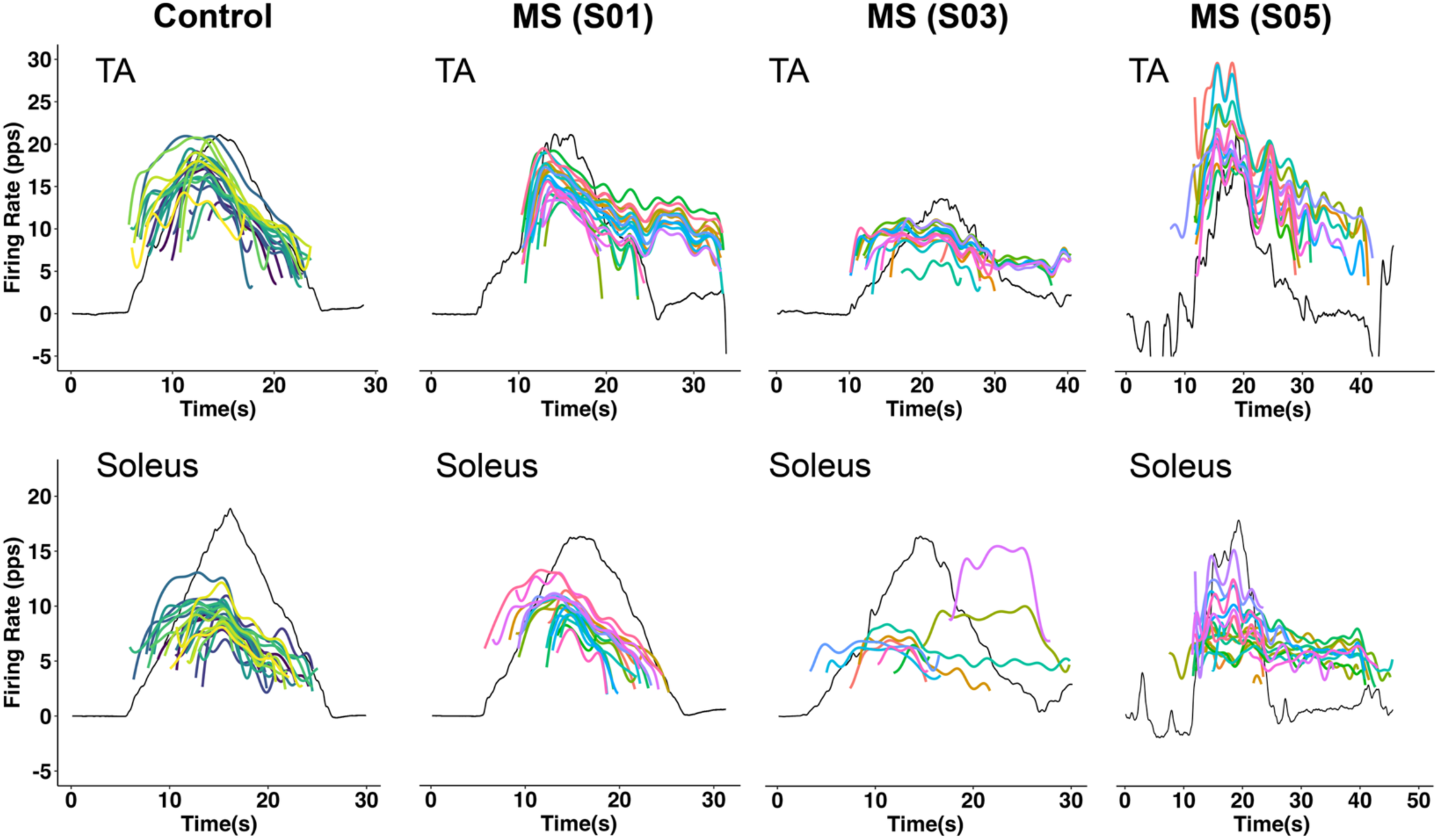
Motor unit firing patterns from single trials. Smoothed motor unit discharge rates from one control participant (left) and three MS participants. Each colored line shows the firing pattern of a different motor unit, and torque over time is shown in black. Data from the TA during dorsiflexion are shown on the top row, and data from the soleus during plantarflexion are shown on the bottom row.

S03 was an older adult (age between 61-65 yo) with mild hemiparesis and spasticity of the right lower extremity. Their motor units changed very little in their rates following a shallow initial acceleration, with self-sustained firing of some of the motor units after task cessation.

S05 had mild bilateral weakness, left lower extremity spasticity, bilateral lower extremity spasms, and central neuropathic pain symptoms (age between 41-45 yo). It was difficult for them to control their torque purposefully at times, and even the anticipation of the impending triangle task caused their ankle muscles to contract unpredictably in a manner that did not occur between trials (see baseline deviations prior to triangle contraction). Their motor units fired at high rates with large acceleration at initial recruitment and notable self-sustained firing after task cessation.

### 3.4. Differences in group means

Figure 4 illustrates MS and control group distribution shapes for the excitation, inhibition, neuromodulation, and firing rate composite variables, shown as probability density functions. Figure 5 shows individual participant values for the reverse engineering features and firing rate characteristics broken down by Group and Muscle.

**Figure 4.**
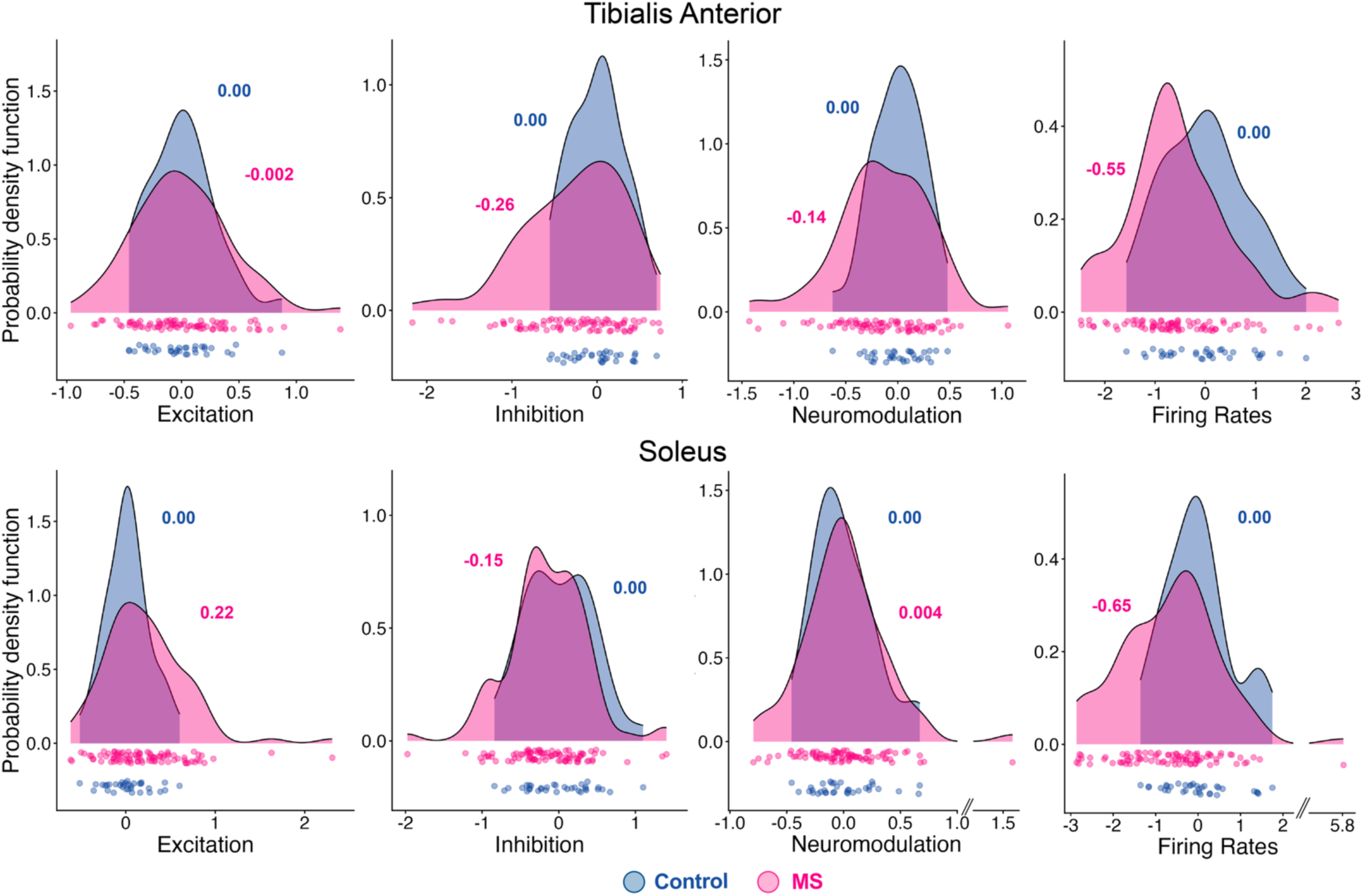
Group distribution shapes for the excitation, inhibition, neuromodulation, and firing rate composite variables. Probability density functions are shown for the control (blue) and MS (pink) groups for the TA (top) and soleus (bottom). The individual participant median values that make up the density functions are shown at the bottom of each plot, colored by group. Estimated marginal means (i.e., adjusted for covariates) are reported in the text.

**Figure 5.**
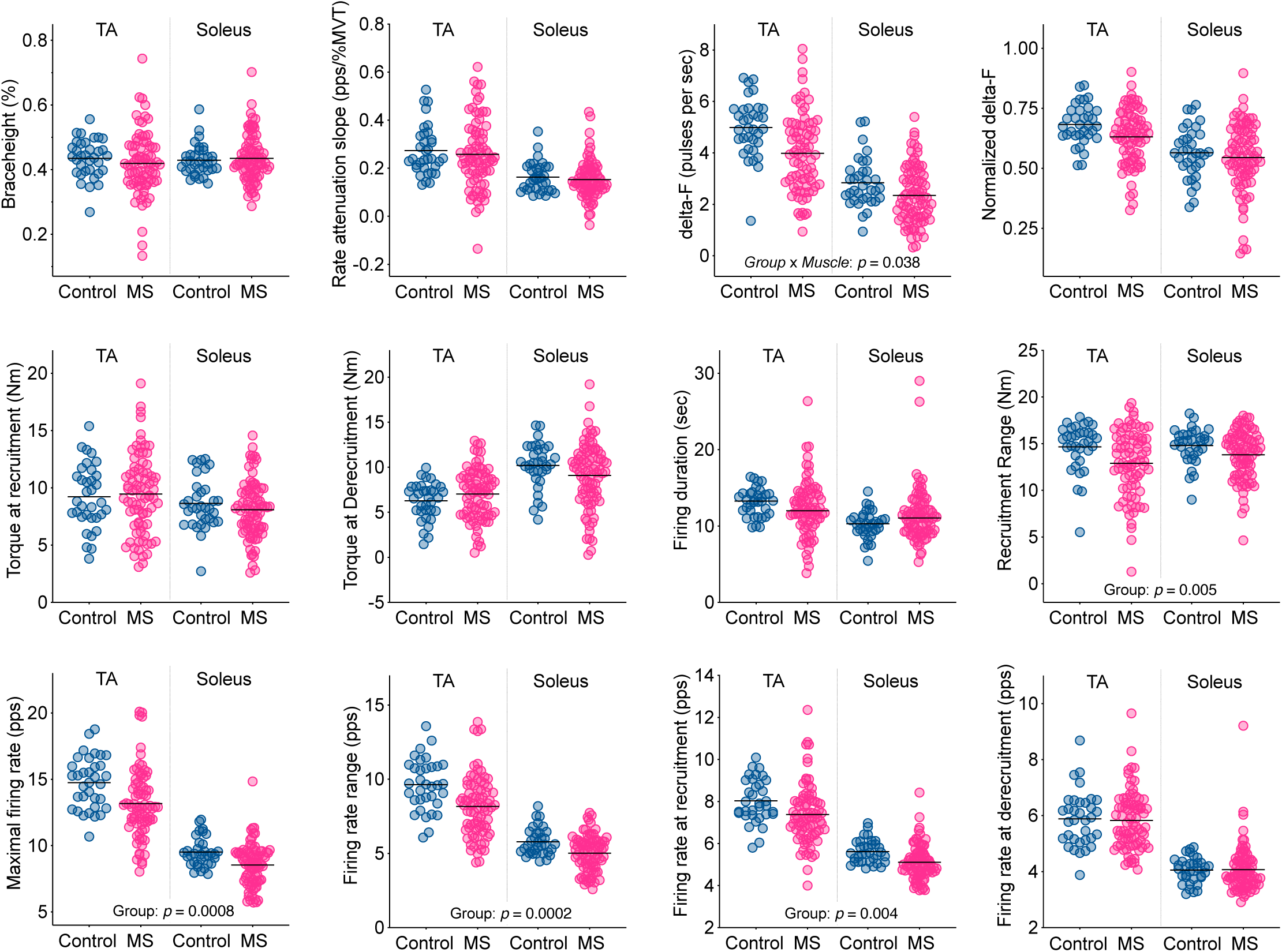
Summary of reverse engineering feature data. Median values for control participants are shown as blue circles and those for MS participants are shown as pink circles. Group mean values for each *Group* and *Muscle* combination are indicated by horizontal black lines. For parameters with a significant effect of Group or Group x Muscle interaction, the associated p-value is displayed on the plot.

We first tested whether the MS and control groups differed on the composite variables and individual reverse engineering features in terms of their group means. For the composite variables, there were significant main effects of *Group* for firing rates (EMM: −0.72 vs. −0.12, *p* = 0.002) and inhibition (EMM: −0.19 vs. −0.02, *p* = 0.048), with the MS group mean was lower for both variables.

There was also a significant *Group* x *Muscle* interaction for the excitation composite (*p* = 0.043), with the soleus mean for the MS group significantly higher than that of controls (EMM: 0.21 vs. −0.001, *p* = 0.01) and significantly higher than the TA mean for the MS group (EMM: 0.21 vs. −0.003, *p* = 0.0002). For neuromodulation, the EMM for the TA and soleus were −0.14 and 0.001 for the MS group and −0.02 and −0.02 for the control group (main effect of *Group: p* = 0.38; *Group* x *Muscle* interaction: *p* = 0.11).

There was a significant main effect of *Group* for five of the 12 individual features. Delta-F (*p* = 0.012), firing rate at recruitment (*p* = 0.004), firing rate range (*p* = 0.0002), maximal firing rate (*p* = 0.0008), and recruitment range (*p* = 0.005), were all lower in the MS group compared with the Control group (mean differences in **Appendix 1 – Table A1**). In other words, on average, motor units from participants in the MS group were recruited at a lower firing rate and across a narrower range of torque, they reached a lower maximal firing rate and exhibited less overall modulation of the firing rate, and they demonstrated less recruitment hysteresis. In addition, there was a significant *Group* x *Muscle* interaction for delta-F (*p* = 0.038), with the corresponding post-hoc test of simple effects indicating that the difference in delta-F between groups was significant for the TA (estimated marginal means (EMM): 4.03 vs. 4.79 pps, *p* = 0.005) but not the soleus (EMM: 2.37 vs. 2.64 pps (EMM), *p* = 0.12).

A complete summary of the linear mixed models results for the individual and composite variables is shown in Appendix 1 – Tables A1, A2, **and** A3.

### 3.5. Group differences in distribution spread and shape

As can be seen in Figure 4, for most of the composite variables, there were extreme MS values at both high and low ends, with the remaining MS values spread more evenly along the range compared to control values which tended to be clustered in the middle of the range. Similarly, in Figure 5, the MS samples were generally more variable than the control samples across all variables, and MS values for many of the variables occurred at both high and low extremes. For all individual variables for both muscles, the MS sample exhibited a wider range than controls (by approximately 1.2 – 3.8x) and a higher coefficient of variation (by approximately 1.1 – 2.1 x).

The variance for many of the composite variables was higher for the MS group compared with controls. For the TA, the MS group had significantly higher variance for the excitation (0.17 vs. 0.09, *p* = 0.048), inhibition (0.35 vs. 0.10, *p* = 0.002), and neuromodulation (0.19 vs. 0.06, *p* = 0.004) composites but not the firing rate composite (1.07 vs. 0.72, *p* = 0.51).

For the soleus, the variance for the MS group was significantly higher for the excitation (0.21 vs. 0.08, *p* = 0.02) and firing rate composites (1.59 vs. 0.62, *p* = 0.044), but not the inhibition (0.27 vs. 0.19, *p* = 0.68) or neuromodulation (0.13 vs. 0.07, *p* = 0.30) composites. These results persisted after excluding the extremely high isolated MS values (Figure 4) and repeating the tests.

The shapes of the mean-centered probability density functions (not shown graphically) were significantly different between groups for all four composite variables for both muscles (TA: Kolmogorov-Smirnov D ranging from 0.29 to 0.55 and *p* ranging from 0.009 to < 0.0001; soleus: Kolmogorov-Smirnov D ranging from 0.37 to 0.63 and all *p* values < 0.00015). The group differences for the soleus persisted when excluding the extremely high isolated MS values (Figure 5) and repeating the tests, with the exception of neuromodulation (D = 0.23, *p* = 0.08).

For the individual variables, the variance of the MS group was significantly higher for the TA for braceheight (0.009 vs. 0.003, *p* = 0.04; Levene’s test), delta-F (2.3 vs. 1.3, *p =* 0.026), torque at de-recruitment (8.1 vs. 4.2, *p* = 0.027), duration (13.2 vs. 3.3, *p* = 0.01), and recruitment range (12.8 vs. 6.6, *p* = 0.016). For the soleus, the variance of the MS group was significantly higher for braceheight (0.005 vs. 0.002, *p* = 0.034), torque at de-recruitment (12.9 vs. 6.3, *p* = 0.034), recruitment range (6.8 vs. 3.6, *p* = 0.032), and maximum firing rate (2.4 vs. 1.2, *p* = 0.037).

Complete results of tests for equality of variance and differences in distribution shape are presented in **Appendix 1 – Table A4** and **Table A5**.

## 4. Discussion

### 4.1. Voluntary motor commands among patients with MS are abnormal in a variety of ways, with some consistencies

Our primary finding is that excitatory, inhibitory, and neuromodulatory components of the motor command are disrupted in a variety of ways among individuals with MS. Specifically, extreme MS values on both ends of the distributions of neuromodulation, excitation, and inhibition indicate different types of disruptions to the motor command in MS, not only when compared to controls, but also within the MS group itself. This pattern is compatible with the heterogeneity of lesion locations and clinical symptoms observed in MS and is fundamentally different than the *relative* consistency of motor pathophysiology among patients with other neurological conditions such as stroke or spinal cord injury.

There were, however, some cases for which disruptions in MS were comparatively consistent. MS values for the inhibition composite were generally shifted lower, particularly for the TA. For the soleus, MS values for the excitation composite were higher. In addition, firing rate characteristics were lower on average in MS for both muscles, which is consistent with previous smaller studies (Rosenfalck & Andreassen, 1980; Rice *et al*., 1992; Bruneau *et al*., 2025). However, despite the *lower* MS group mean values, we also found several extreme MS values that were *higher* than any control values. These were notable exceptions that seem to represent more than typical population variability.

In subsequent sections, we interpret these high and low values and discuss how the findings provide insight into possible mechanisms underlying the range of motor deficits among patients with MS.

### 4.2. Possible scenarios resulting in high or low neuromodulation in MS

To the best of our knowledge, the function of descending monoaminergic neuromodulatory pathways in humans with MS has not previously been investigated prior to this study. Our finding that neuromodulatory input to motoneurons in people with MS can occur across a strikingly wide range with both high and low extremes is a valuable insight into how MS degrades motor function in different patients. It is in stark contrast to the uniformly elevated PIC amplitudes that have been previously found in hemiparetic stroke and spinal cord injury, neurological populations with extensive white matter injury that are the most similar to MS.

From previous work in these populations, we know of two potential scenarios by which *increased* neuromodulation can occur. First, with supraspinal corticospinal injury from hemiparetic stroke, the reticulospinal tract has an increased influence on motor output as a result of cortico-reticular disinhibition and serves as compensatory mechanism to generate movement in light of corticospinal injury (McPherson *et al*., 2018*a*; Karbasforoushan *et al*., 2019; McPherson & Dewald, 2022; Mooney *et al*., 2025). In addition to its excitatory fibers, the reticulospinal pathway includes the raphespinal and ceruleospinal projections that deliver monoaminergic neuromodulatory input to motoneurons (Veasey *et al*., 1995; Schwarz *et al*., 2008; Heckman *et al*., 2009; McPherson *et al*., 2018*b*). In MS, a similar process could occur following demyelination and axonal loss within the corticospinal pathway.

Second, spinal cord injury also leads to increased neuromodulation of motor unit discharge, but in a paradoxical manner. With spinal injury, the raphespinal and ceruleospinal tracts can be disrupted or severed, drastically limiting or preventing the delivery of monoaminergic neuromodulatory input to motoneurons and causing profoundly unexcitable motoneurons (Hounsgaard *et al*., 1988; Harvey *et al*., 2006). However, over time, the lack of serotonin and norepinephrine leads paradoxically to an *increase* in PIC amplitude, mimicking the effects of neuromodulatory input. This occurs from upregulation of constitutively active serotonin (5-HT_2c_) and norepinephrine receptors (⍺_1_) on motoneuron dendrites that facilitate PICs in the absence of the neurotransmitters (Murray *et al*., 2010; Rank *et al*., 2011; Tysseling *et al*., 2017; Mahrous *et al*., 2023). In MS, there is strong evidence from animal models that disruption of descending NM pathways is likely (see next section), which could lead to a similar process for restoring PICs, resulting in high values of neuromodulation as assessed with the reverse engineering model.

By extension, our findings of *low* neuromodulation in MS could indicate disruption to descending neuromodulatory pathways *without* compensatory upregulation of serotonin and norepinephrine receptors. Restoration of PICs through this process is vital for increasing voluntary muscle activation after spinal cord injury (Murray *et al*., 2010), so its absence in some patients with MS could have a profound impact on muscle strength.

### 4.3. Evidence for disruption to the descending monoaminergic neuromodulatory system in MS

Because demyelination is a primary mechanism of CNS injury in MS, our findings of altered neuromodulation are perhaps surprising given that descending neuromodulatory pathways have little-to-no myelination.

However, findings from animal models and study of *ascending* monoaminergic projections in humans provide compelling evidence to support the likelihood of their disruption.

A series of studies by White and others (1970s-1990s), using rodent models of MS (rats and guinea pigs with experimental autoimmune encephalomyelitis (EAE)), found morphological damage to descending monoaminergic axons as they encountered blood vessels with perivascular inflammatory cuffs (Bieger & White, 1981; White *et al*., 1985, 1989, 1990; White & Bowker, 1988; Vyas *et al*., 1988). They also found reduced levels of serotonin and norepinephrine in the spinal gray matter (Lycke & Roos, 1973; White *et al*., 1983, 1990; Krenger *et al*., 1986; Samathanam *et al*., 1991) and reduced serotonin receptor sensitivity in the spinal cord (White, 1979; White & Bieger, 1980). These changes seem to be specific to monoaminergic pathways, which could be more susceptible to injury because of their small diameter and thin or lack of myelination (White & Bowker, 1988; Polak *et al*., 2011).

Importantly, this body of work demonstrated that the observed changes to the descending monoaminergic system were often associated with clinical symptoms, in terms of their location (lower extremity > upper extremity (White *et al*., 1983, 1985, 1989; Krenger *et al*., 1986, 1989), time course (appearing with the earliest motor symptoms (White *et al*., 1989, 1990)), and severity (correlated with extent of hindlimb paralysis (White *et al*., 1990)). Additionally, preclinical interventions in mice with EAE that increased monoamines in the spinal cord via electrical stimulation of the nucleus raphe magnus (Madsen *et al*., 2017), or chemogenetic activation of the noradrenergic neurons in the locus ceruleous (Torrillas-de La Cal *et al*., 2023), led to substantially improved motor function. Although these improvements can be attributed, at least in part, to the widespread anti-inflammatory and neuroprotective effects of these neurotransmitters, their ability to restore neuromodulatory action on motoneurons undoubtedly contributed.

Finally, in further support of alterations to the monoaminergic system in MS, *ascending* noradrenergic, serotonergic, dopaminergic projections from the brainstem to the brain have been studied with regard to prevalent clinical symptoms of fatigue, depression, and cognitive dysfunction (Feinstein *et al*., 2016; Manjaly *et al*., 2019; Carandini *et al*., 2021*a*). Collectively, findings from studies using mice with EAE and patients with MS (Gadea *et al*., 2004; Polak *et al*., 2011; Carandini *et al*., 2021*c*, 2021*b*) indicate that ascending monoaminergic transmission is less effective due to both structural damage and an inflammation-induced reduction in monoamine synthesis (Manjaly *et al*., 2019; Carandini *et al*., 2021*a*). Some studies have shown relationships among various measures of ascending monoaminergic pathway structure and/or function with clinical symptoms of fatigue (Carandini *et al*., 2021*c*), depressive symptoms (Carotenuto *et al*., 2020), and cognitive dysfunction (Gadea *et al*., 2004; Carotenuto *et al*., 2020).

Taken together, our findings and the above body of work highlight the intriguing possibility that disruptions to the descending monoaminergic neuromodulatory system contribute to motor deficits in MS, which warrants further study.

### 4.4. Implications of abnormal inhibition and excitation on force generation

Excitatory and inhibitory inputs to motoneurons strongly influence the two primary mechanisms for the CNS to increase muscle force. The pattern of inhibitory input can limit or accentuate rate modulation, and the distribution of excitatory input affects the time course and efficiency of orderly recruitment.

Our excitation composite findings show that abnormal recruitment of motoneurons occurs in different ways among patients with MS. Higher excitation composite values, as we saw with the TA for some participants with MS and more consistently with the soleus, indicate relatively more excitatory input to larger motoneurons compared with controls. This distribution lowers the recruitment threshold for those motoneurons and compresses the overall recruitment range. As a result, these participants may have difficulty with slow, graded contractions because more motoneurons are recruited over a shorter range of synaptic drive. On the other hand, lower excitation composite values, as we saw in the TA for some participants with MS, indicate relatively more excitatory input to smaller motoneurons compared with controls. This distribution widens the overall recruitment range, and as a result, these participants may have difficulty with fast and forceful movements.

Characterizing the distribution of excitatory input using the reverse engineering paradigm also provides insight into which source(s) of excitatory input are disrupted in MS. Voluntary skilled movement is controlled by excitatory inputs to motoneurons from multiple sources (e.g., descending, afferent) that are distributed non-uniformly among small and large motoneurons in different ways (Powers & Binder, 2001; Binder *et al*., 2002). Descending motor pathways (e.g., corticospinal, vestibulospinal) provide more excitatory current to larger motoneurons, whereas Ia sensory afferents that deliver crucial proprioceptive information provide more excitatory current to smaller motoneurons (Heckman & Binder, 1988, 1993). In the future, use of the reverse engineering model along with other neurophysiological approaches (e.g., stimulation of Ia afferents) in the same patients can help elucidate how different sources of excitatory input are disrupted.

Our findings also demonstrated that in general, patterns of inhibitory input were closer to the balanced inhibition extreme compared with controls. Accordingly, motor unit rate modulation was reduced for many participants with MS (see Figure 4, rate attenuation slope and firing rate range). Given the importance of rate modulation for increasing muscle force (Binder *et al*., 2011; Heckman & Enoka, 2012), abnormal balanced inhibition could be an important driver of strength deficits.

### 4.5. Limitations

Some limitations in the extent to which experimental data from clinical populations can be interpreted within the reverse engineering framework should be considered. For example, it is possible that the characteristics of the actual excitatory, inhibitory, and neuromodulatory inputs in some patients could be outside the range over which they were simulated by Chardon et al. (2024), which would lead to less certainty in the validity and interpretation of the reverse engineering features. Fortunately, the input parameter ranges the authors used are sufficiently wide to support our overall finding that motor commands are disrupted in different ways in MS, even if the most extreme data points were to result from true input values that are outside of those simulated. Additionally, as the reverse engineering approach is further developed to model the effect of other parameters on motor unit firing patterns (e.g., level of tonic inhibition, after-hyperpolarization duration, PIC voltage threshold, time course of excitation, etc.), we will be able to characterize abnormal excitatory, inhibitory, and neuromodulatory inputs to motoneurons in clinical populations more precisely.

The differences in maximal dorsiflexion and plantarflexion strength between MS and control groups (see **Appendix 1 – Figure A1**) could also influence some of the reverse engineering features. For example, participants with MS who were weaker than controls may not have recruited as many larger motoneurons during the 20% MVT task compared with controls, which could affect features related to recruitment threshold. However, several aspects of the dataset indicate that between group differences cannot be attributed solely to strength differences. First, there were many participants with MS with maximal strength within normal limits who demonstrated abnormal values for the composite variables and reverse engineering features. Second, we controlled for torque at recruitment in the between group comparisons for the reverse engineering features and still found differences in means and variability. Finally, when repeating the between group comparisons for the composite variables and including a covariate of maximal torque (**Appendix 1 – Table A3**), the results were generally unchanged, but the magnitude of the group mean differences increased and the main effect of *Group* for the neuromodulation became significant.

Finally, we should consider whether any medications taken by MS participants could contribute to the group’s varied differences from controls. We explored qualitatively the composite variable values for participants with MS taking anti-spastic medications (e.g., baclofen, tizanidine) as well as monoamine reuptake inhibitors. We observed no obvious systematic effects of the medications, in that patients taking them were spread throughout the range of values. Additionally, we will consider the effects of different disease characteristics (e.g., disease duration, disease subtype) on our findings in future analyses. Such characteristics are typically correlated with the level of motor impairment, and thus they will require a dedicated manuscript to disentangle any effects they may have.

### 4.6. Summary and Future Work

Our findings that excitatory, inhibitory, and neuromodulatory components of the motor command are disrupted in a variety of ways among patients with MS provide a solid basis for explicating the nature of these disruptions. For example, follow up investigations will explore whether this variation occurs at the individual patient level or if there are pathophysiological subgroups within MS. Further, determining whether values of the composite variables relate to measures of motor and functional impairment in MS will provide valuable insight into the underlying pathophysiological mechanisms and how they vary among patients.

Additionally, our study demonstrates that the reverse engineering framework of Chardon et al. (2024) can be feasibly and meaningfully applied to human data to reveal new insights that were otherwise inaccessible. Our success in to capturing many different types of abnormal motor commands in MS suggests that the framework is likely to be of value with a variety of other clinical populations.

## 5. Methods

### 5.1. Ethical Approval

All participants provided written informed consent for participation in the study, which was approved by the Institutional Review Board of Washington University and conformed to the standards set by the Declaration of Helsinki, except for registration in a database (Institutional Review Board protocol numbers: 202209111 and 202404145 (MS participants); 202009146 and 202211021 (control participants without neurological injury or disease).

### 5.2. Participants

Participants with MS were recruited from the John L. Trotter Multiple Sclerosis Center at Washington University and/or the Washington University Physical Therapy outpatient clinic. Individuals without neurological injury or disease (controls) were recruited through the Washington University Research Participant Registry or by word of mouth. They were included in the study if their sex and age (within 5 years) matched those of an MS participant.

Potential participants were screened prior to study enrollment. Participants with MS were required to have (1) a confirmed diagnosis of MS made by a neurologist, (2) the ability to generate a muscle contraction in at least one of the tested muscles for ≥ 20 sec, (3) no clinical relapses within the last six months, (4) the ability to transfer into the testing setup with physical therapist assistance if needed, and (5) sufficient passive range of motion in the tested leg to sit comfortably in the testing setup.

Participants from both groups were excluded if they (1) had a history of severe musculoskeletal or peripheral neurological injury to the tested leg, (2) pain or low skin integrity that would prevent sitting in the testing chair, (3) severe cognitive or visual deficits, (4) uncontrolled hypertension or (5) central neurological injury or disease (other than MS for the MS group).

Ninety-two individuals with MS met all enrollment criteria and completed the study. Data could not be used for 3 participants because of profound weakness in both dorsiflexion and plantarflexion that prevented completion of the task (N = 1), discomfort during testing that precluded completion of the experiment (N = 1), and because no usable motor units could be obtained from either muscle (N = 1). Therefore, data from 89 participants were included in the analysis (68 females, 21 males; mean age ± SD: 51.4 ± 10.6 years, range 29-79; mean symptom duration ± SD: 16.8 ± 10.5 years, range: <1 - 48; **Table 2**). Sixty-three participants had relapsing remitting MS, 16 had secondary progressive MS, and 10 had primary progressive MS. Participant disability was assessed using the Expanded Disease Disability Scale (EDSS), and enrolled participants had scores ranging from 0 (no disability) to 7 (severe disability) (mean ± SD: 2.9 ± 1.9). The mean ± SD time to complete a forward 25-ft walk was 7.7 ± 6.9 sec (range 3.72 – 54.0), and the mean ± SD time to complete a backward 25-ft walk (Edwards *et al*., 2020) was 14.8 ± 16.3 sec (range 3.52 – 122.2). Based on clinical MRI, participants had lesions in the following general locations (listed as percentage of participants): cortical and/or juxtacortical (90%), periventricular (97%), callosal (78%), subcortical (26%), brainstem (51%), cerebellar (56 %), cervical spinal cord (79%), and thoracic spinal cord (55%) (**Table 2**). Seventy-eight of the 89 participants were on disease-modifying medications, and thirty-four of the 89 participants were on an anti-spastic medication.

**Table 2.**
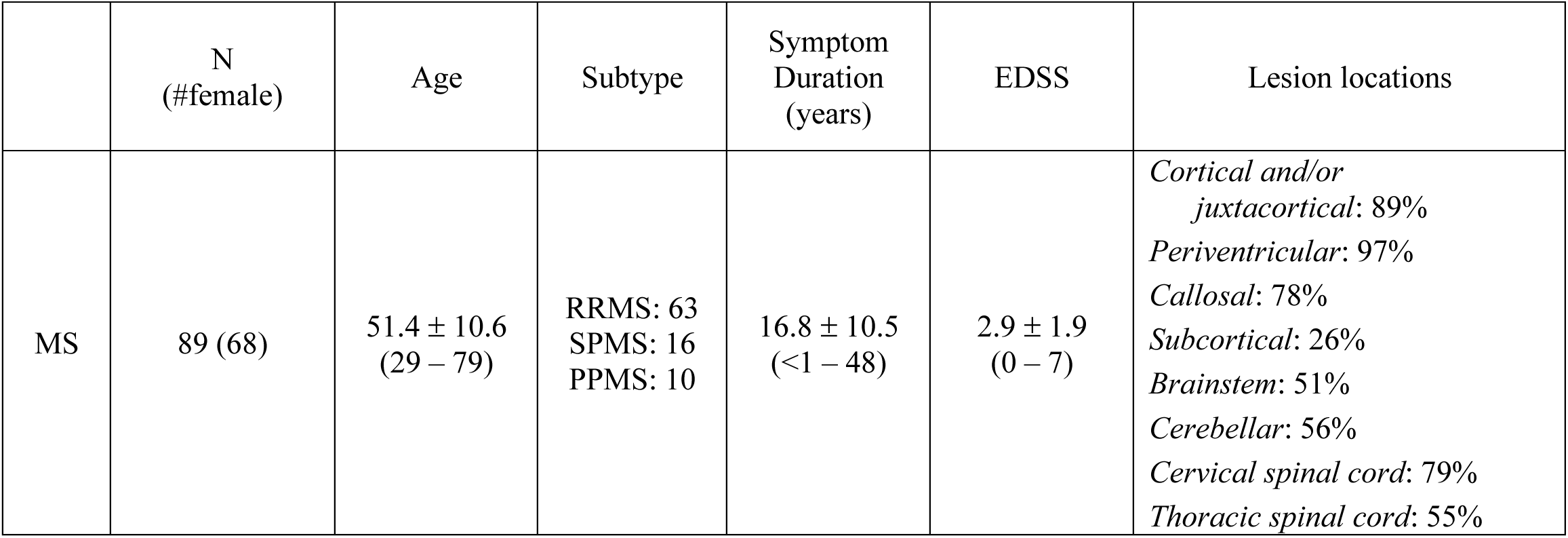

|  | N<br>(#female) | Age | Subtype | Symptom<br>Duration<br>(years) | EDSS | Lesion locations |
| --- | --- | --- | --- | --- | --- | --- |
| MS | 89 (68) | $51.4 \pm 10.6$<br>(29 – 79) | RRMS: 63<br>SPMS: 16<br>PPMS: 10 | $16.8 \pm 10.5$<br>($<1 - 48$) | $2.9 \pm 1.9$<br>(0 – 7) | *Cortical and/or*<br>*juxtacortical: 89%*<br>*Periventricular: 97%*<br>*Callosal: 78%*<br>*Subcortical: 26%*<br>*Brainstem: 51%*<br>*Cerebellar: 56%*<br>*Cervical spinal cord: 79%*<br>*Thoracic spinal cord: 55%* |

Thirty-four individuals without neurological injury met all enrollment criteria and completed the study (22 females, 12 males; mean age ± SD: 45.5 ± 14.2 years, range 25-75). For each participant with MS, there was an average of four controls in the dataset who were of matched age and sex (range 1-9).

### 5.3. Experimental Setup and Data Collection

Experimental testing was conducted using the Biodex System 4 Pro dynamometer (Biodex Medical Systems, Shirley, NY) (Figure 2) to measure isometric joint torque. Participants sat upright in the experimental chair with their tested foot secured to the ankle attachment of the dynamometer. Their leg was positioned at 90° hip flexion and 10° ankle plantarflexion, which resulted in approximately 20° knee flexion. A TV monitor displayed real-time visual feedback of torque data.

High-density surface EMG was recorded in monopolar fashion from 64-channel rectangular electrode grids with 8 mm interelectrode distance (GR08MM1305, OT Bioelettronica, Inc.) secured to skin that had been prepared using abrasive paste. One electrode grid was placed over the tibialis anterior muscle belly, and two electrode grids were placed over the soleus muscle belly, one on each side of the Achilles tendon. Care was taken to ensure the electrode remained on the soleus and did not cover the lateral ankle musculature. Analog EMG and torque signals were acquired using the Quattrocento data acquisition system (OT Bioelettronica, Inc.) with a sampling rate of 2048 Hz, high-pass filter of 10 Hz, and low-pass filter of 900 Hz.

### 5.4. Experimental Protocol

Participants completed the protocol with both legs, performed in randomized order. In this analysis, we include data from the MS participants’ more impaired leg and from the control participants’ dominant leg. We included data from their dominant leg (1) because preliminary data in our lab suggest no meaningful differences in our measures between legs in controls and (2) as a matter of efficiency because the time-consuming decomposition and analysis were already ongoing as part of a separate study. The MS participants’ more impaired leg was determined based on patient report of motor symptoms. For the participants who did not have appreciable motor symptoms, the more impaired leg was determined based on sidedness of non-motor symptoms or was chosen randomly.

First, each participant’s isometric maximum voluntary torques (MVTs) were measured in dorsiflexion and plantarflexion, performed in randomized order. Trials within a direction were repeated until three trials within 85% of the maximum value were obtained, without the last one being the highest. Participants were given vigorous verbal encouragement during MVT trials and several minutes of rest in between trials.

Then, participants performed submaximal triangular contractions with a visual template provided on the screen (blue line in Figure 1). They were instructed to rest for 5 sec at trial initiation, then slowly increase their isometric torque at a rate of 2% MVT/sec over 10 sec to a peak of 20% MVT, and then slowly decrease their isometric torque at the same rate prior to resting for another 5 sec. Trials were repeated with rest breaks in between until the participant completed three to five trials with appropriately smooth and triangular torque traces (typically 5-10 total per direction). The triangular contractions were performed in both dorsiflexion and plantarflexion, with the trial blocks for each direction completed in randomized order. An experienced operator carefully identified torque traces that were acceptable for subsequent analysis based on visual inspection of smoothness, symmetry, and a lack of influence of any deviations thereof on our reverse engineering parameters (Hassan *et al*., 2020).

### 5.5. Data Analysis

#### 5.1.1. Motor unit decomposition and processing of motor unit spike trains and torque signals

Offline, high-density surface EMG channels were visually inspected, and channels with substantial noise and/or movement artifacts were removed from further analysis. The remaining channels were decomposed automatically into individual motor unit spike trains using a convolutive blind source separation algorithm with a silhouette threshold of 0.85 (Negro *et al*., 2016). The accuracy of each resulting spike train was inspected by a trained team member and improved iteratively using a well-validated local re-optimization technique implemented using a custom MATLAB (RRID: SCR_001622) graphical user interface, to correct minor errors from the automatic decomposition or discard the motor unit (Del Vecchio *et al*., 2020; Hug *et al*., 2021; Hassan *et al*., 2021; Jenz *et al*., 2023). Motor units were tracked across trials using cross-correlation of their motor unit action potential profiles (Martinez-Valdes *et al*., 2017; Thompson *et al*., 2018).

A custom MATLAB (RRID: SCR_001622) program was used to define a motor unit’s recruitment time as the first spike time to be followed by sustained firing with subsequent inter-spike intervals with a duration less than 500 ms. A motor unit’s de-recruitment time was defined as the last spike time before an inter-spike interval of greater than 500 ms.

We used a support vector machine regression model with kernel value of 2.2 to predict smoothed motor unit discharge profiles from instantaneous discharge rates with accurate onset and offset discharge rates and low levels of residual error (Beauchamp *et al*., 2022). Torque signals were smoothed (acausal one-sided moving average filter with window length of 250 ms), baseline corrected, and normalized to the MVT value for that torque direction.

#### 5.5.1. Motor unit decomposition performance

In total, 19,491 decomposed motor unit spike train recordings were suitable for analysis, with an average of 17.6 per trial for the TA and 16.5 per trial for the soleus. Of these, 12,340 were designated as recordings from unique motor units, with an average number of unique motor units per participant of 41.8 for the TA and 60.2 for the soleus. The total number of spike train recordings and average number of spike trains per trial for each muscle, sex, and group combination are reported in **Table 3** along with similar descriptive statistics for the number of unique motor units per participant.

**Table 3.** The mean ± SD (total) number of motor unit spike trains decomposed per trial (top) and the mean ± SD (total) number of unique motor units per participant (bottom) for each group, muscle, and sex combination.

|  | *mean ± SD (total) number of motor unit spike trains decomposed per trial* |  |  |  |
| --- | --- | --- | --- | --- |
|  | **MS** |  | **Control** |  |
|  | *Female (N = 68)* | *Male (N = 21)* | *Female (N = 22)* | *Male (N = 12)* |
| **TA** | 15.5* ± 8.3 (4570) | 23.1* ± 9.4 (2099) | 15.7 ± 5.8 (1556) | 26.0 ± 8.1 (1353) |
| **Soleus** | 14.5 ± 8.3 (4564) | 22.7 ± 11.8 (2167) | 16.1 ± 6.6 (1767) | 22.0 ± 10.3 (1415) |
|  | *mean ± SD (total) number of unique motor units per participant* |  |  |  |
|  | **MS** |  | **Control** |  |
|  | *Female (N = 68)* | *Male (N = 21)* | *Female (N = 22)* | *Male (N = 12)* |
| **TA** | 35.2* ± 22.2 (2253) | 57.3* ± 29.6 (1146) | 37.5 ± 17.8 (824) | 59.3 ± 26.9 (712) |
| **Soleus** | 47.9 ± 30.7 (3257) | 74.8 ± 37.8 (1571) | 65.6 ± 36.3 (1444) | 94.4 ± 44.1 (1133) |

\*There were four females and one male in the MS group that did not have usable motor unit recordings for the TA. For three participants, this was due to an inability to complete the task (see Section 3.3), and for the other two, it was because of poor decomposition results. Those participants are not included in the aggregate numbers for the TA.

#### 5.5.2. Calculation of reverse engineering features

For each smoothed motor unit firing profile, we calculated eight reverse engineering features (braceheight, rate attenuation slope, normalized delta-F, torque at recruitment, torque at de-recruitment, firing duration, and recruitment range) as well as four firing rate characteristics (firing rate at recruitment, firing rate at de-recruitment, firing rate range, maximal firing rate) (Figure 2A**, 2B**).

Torque at recruitment and de-recruitment were calculated as the torque (%MVT) produced at the time of the first and last instantaneous discharge rate, respectively. Firing duration (sec) was calculated as the difference between the time of motor unit de-recruitment and motor unit recruitment. Recruitment range (%MVT) was calculated as the difference in torque produced when the last motor unit was recruited and the torque produced when the first motor unit was recruited.

Delta-F (pps) was calculated to quantify recruitment hysteresis using the paired motor unit technique (Gorassini *et al*., 2002), which is the most established method for estimating PIC prolongation in humans (Mesquita *et al*., 2024) and has been validated in animal models and using motoneuron simulations (Powers & Heckman, 2015). Motor units within a trial were compared in a pairwise fashion, calculating delta-F for the higher threshold motor unit as the difference in the smoothed firing rate of the lower threshold motor unit between the times of recruitment and de-recruitment of the higher threshold motor unit (Figure 2A). We used the following criteria to identify appropriate motor unit pairs (Gorassini *et al*., 2002; Stephenson JL & Maluf KS, 2011; Hassan *et al*., 2021; Jenz *et al*., 2023): 1) recruitment time difference of > 1 sec, 2) correlation of the smoothed discharge rates with R^2^ > 0.7, and 3) rate modulation of the control unit that is at least 0.5 pps greater than the delta-F value.

One critique of the delta-F technique is that the level of rate modulation in the low threshold motor unit may partially confound the measure (e.g., delta-F calculated with a low threshold motor unit with minimal rate modulation could underestimate PIC amplitude). Thus, we also used a normalized delta-F, which is the calculated delta-F divided by the total possible delta-F (using the lower threshold motor unit’s minimum firing rate during the descending portion of the contraction rather than the firing rate at the time of high threshold de-recruitment). For both forms of delta-F, we obtained one value per motor unit by averaging over all values calculated with different lower threshold motor units (Hassan *et al*., 2021; Jenz *et al*., 2023).

The braceheight parameter quantifies the overall non-linearity of increasing motor unit discharge by calculating the extent to which the smoothed firing rate trajectory from recruitment to maximal firing rate deviates from a theoretical linear increase (Beauchamp *et al*., 2023). Braceheight is calculated on the firing rate vs. torque function as the magnitude of the maximal orthogonal distance between the firing rate trajectory and a straight line between recruitment and maximal firing rate (Figure 2B). It is then normalized to the maximum possible braceheight value that would be obtained from a theoretical right triangle with the same start and end points (Beauchamp *et al*., 2023).

Rate attenuation slope (pps/%MVT) is calculated as the linear regression slope for the segment of the firing rate vs. torque function between the inflection point and the maximal firing rate (Beauchamp *et al*., 2023). Some motor units peak quickly after the acceleration phase, complicating calculation of the rate attenuation slope.

Therefore, if the duration of the rate attenuation phase was less than 1 second, we calculated the rate attenuation slope over the firing rate vs. torque function from the inflection point to 1 second prior to the maximal torque.

This allowed for calculation of negative rate attenuation slopes when found.

Firing rate at recruitment and de-recruitment (pps) were calculated as the first and last point of the smoothed firing profile, respectively. Maximal firing rate was calculated as the maximum value of the smoothed firing profile, and firing rate range was calculated as the maximum value minus the minimum value of the smoothed firing rate.

We aggregated values for each reverse engineering feature across motor unit recordings per participant and muscle so that we would have one participant-level value for each muscle. For each motor unit that was identified in multiple trials, we averaged its values across trials to provide one value per feature per motor unit. Then, we calculated the median value across all motor units, resulting in one value per feature per person.

#### 5.5.3. Calculation of composite variables

First, for each muscle, we converted each participant’s median value into a Z-score, calculated relative to the control group mean and standard deviation for that muscle. Then, we calculated the composite score for each component using a weighted average of the reverse engineering features contributing to prediction of that input (Equations 1-3). The weights for each feature were based on the features’ mutual information score with a given component, as determined by Chardon et al. (2024), with signs added to denote the relationships between variables based on prior literature (Powers *et al*., 2012; Beauchamp *et al*., 2023; Chardon *et al*., 2024) (**Table 1**). We also calculated four key firing rate characteristics (maximum firing rate, firing rate range, firing rate at recruitment, and firing rate at de-recruitment) which we averaged to create a firing rate composite score (Equation 4). The equations are as follows:

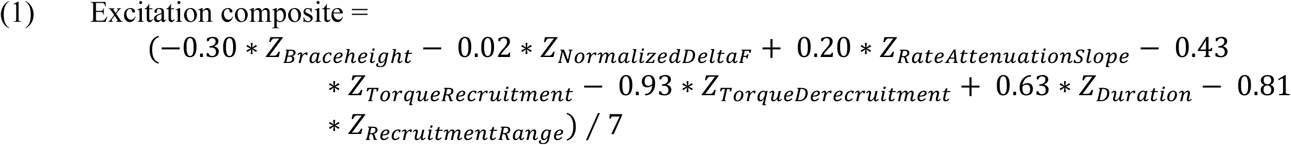

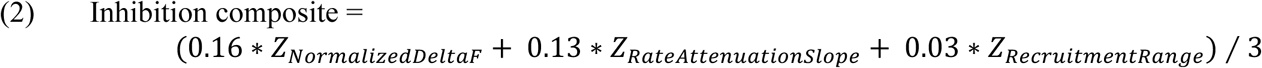

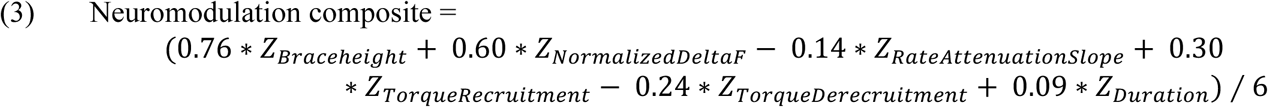

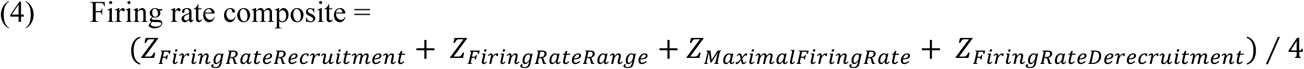

### 5.6. Statistical Analyses

All statistical analyses were performed in R (version 4.4.0, (R Core Team, 2024), RRID:SCR_001905) using RStudio (RRID:SCR_000432) with statistical significance set to ⍺ = 0.05. As described below, we used different linear mixed-effect regression models to address different research questions (Bates *et al*., 2015). Each of these models included specific fixed effects (depending on the tested hypotheses) and random effects (to account for statistical dependencies in how the data were structured; described below and illustrated in **Figure A2**).

#### 5.6.1. Differences in Group Means

To assess between-group differences for each reverse engineering feature, we performed a linear mixed effects model with fixed effects of *Group*, *Muscle*, *Group-by-Muscle*, and covariates of *Sex* and *Age_c* as participant-level predictors. Torque at recruitment was grand-mean centered (*TQ_recrt_c*) and included as a fixed effect, serving as a motor unit-level covariate (Jenz *et al*., 2023) (except for the model with *TQ_recrt* as the outcome measure).

To fully account for the dependencies within the data, we included random intercepts for *Participant, Trial,* and *Motor Unit* within *Muscle* within *Participant* as well as random slopes for *Muscle* within *Participant* and *Muscle* within *Trial* (R syntax: (1+Muscle | Participant) + (1+Muscle | Trial) + (1 | Participant : Muscle : MotorUnit)). For some outcome variables, the random slope of *Muscle* within *Trial* did not account for any estimated variance and was therefore removed from the model to avoid a singularity. For a subset of these variables, the same scenario occurred with the random intercept of *Trial. Group, Muscle*, and *Sex* were contrast coded so that the associated coefficients would correspond to the mean difference between the two levels of each factor (coded as: MS - Control, TA - soleus, Male - Female). The coefficient of the *Group* x *Muscle* interaction is interpreted as the mean difference between groups of the mean difference between muscles.

Estimated mean differences and coefficients are reported along with their 95% confidence intervals.

Visual inspection of Q-Q plots for many of the linear mixed effects models revealed moderate-to-large departures from normality. Therefore, we applied parametric bootstrapping methods to all models using the *pbmodcomp* function from the *pbkrtest* package (Halekoh & Højsgaard, 2014) with 2000 simulations. We used the Bartlett statistic to determine statistical significance of fixed effects (Halekoh & Højsgaard, 2014).

To assess between-group differences for each composite variable, we performed a linear mixed effects model with fixed effects of *Group*, *Muscle*, *Group-by-Muscle*, and covariates of *Sex* and *Age_c* with a random intercept of *Participant* to account for the repeated measure of Muscle (R syntax: (1 | Participant)). Because the composite variables were calculated on the participant level rather than the motor unit level, and because there were no other repeated measures, no additional random effects are necessary. Examining the distribution of model residuals revealed no substantive departures from normality, and therefore, we used the F-test to determine statistical significance of fixed effects.

#### 5.6.2. Group differences in distribution spread and shape

To compare how the MS and control samples were distributed around their means, we examined the shapes and spread of the distributions of participant median values. We tested for equality of variance between groups for each reverse engineering feature/composite variable and muscle using Levene’s test from the *car* package (Fox & Weisberg, 2019). Then, we tested for statistical differences between the overall MS and control distribution shapes for each composite variable by centering the distributions around their group mean and applying the two-tailed two-sample Kolmogorov-Smirnov test (*stats* package included in R).

## 6. Author contributions

All experiments were performed at Washington University School of Medicine in St. Louis, MO.

Conceptualization: LM, KL, RN, and AC

Funding acquisition: LM, KL, RN, and AC

Methodology: LM, KL, DF, JB, FN, RN, AC

Investigation: LM, SS, DF

Data curation: LM, SS, DF

Formal Analysis: LM, KL

Visualization: LM, KL

Writing – original draft: LM

Writing – review and editing: LM, KL, SS, DF, JB, FN, RN, AC

## Data Availability

Data produced in the present study are available upon reasonable request to the authors.

## Acknowledgments

We thank the study participants for their time and effort; the John L. Trotter MS Center at WashU Medicine and the WashU Medicine Physical Therapy Clinics for assistance with recruitment of MS patients; Tanner Reece, MS for assistance with data collection, preliminary analyses, and figure preparation; Abbey Painter, BS for assistance with data collection and figure preparation; CJ Heckman, PhD for discussions about interpretation of findings; and Matthieu Chardon, PhD for providing exact mutual information scores.

## 9. Data Availability

Data from the study will be publicly available to accompany the publication of the final version of this paper.

## 11. Appendix

### 11.1. Differences in maximal strength

**Figure A1.**
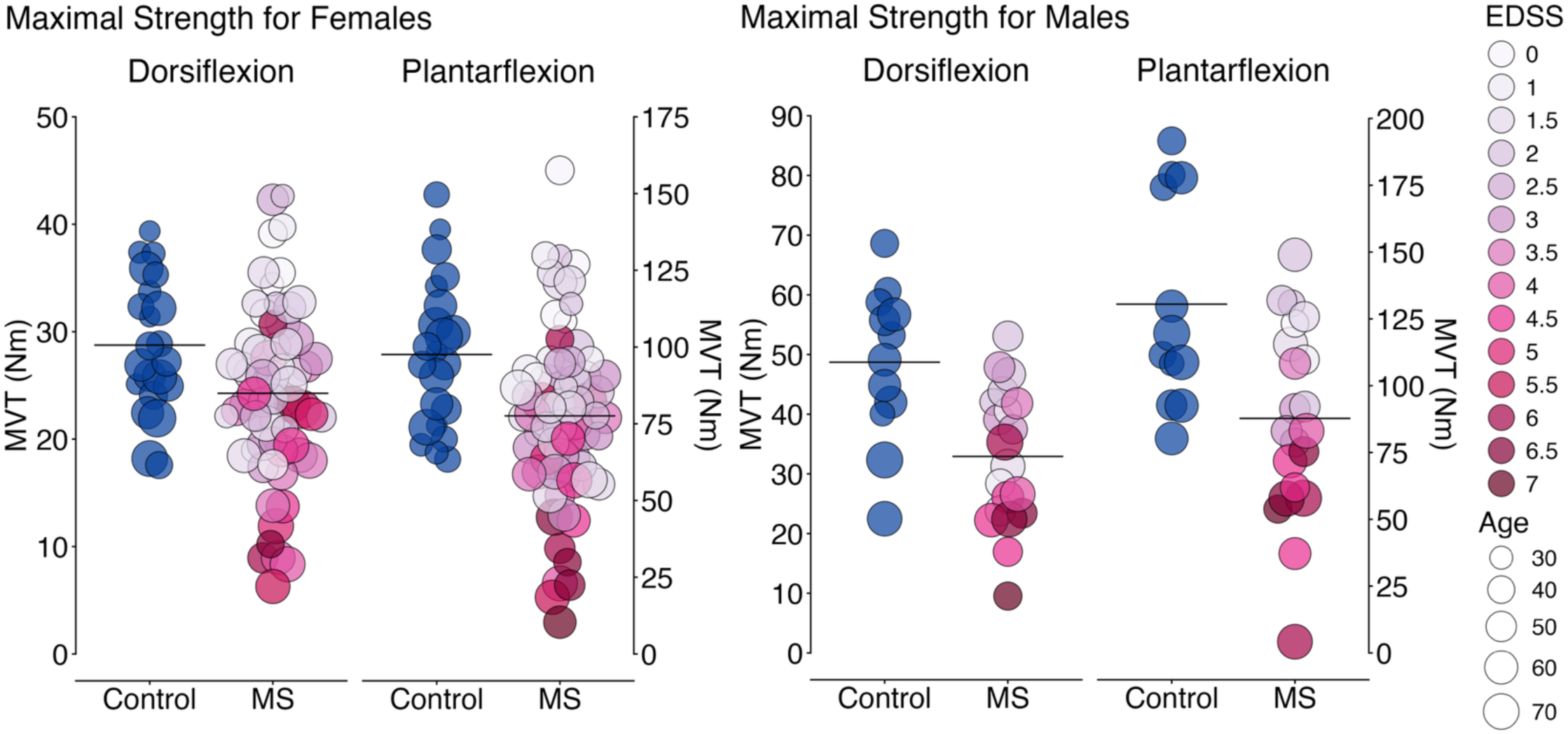
Maximum voluntary torque (MVT) in dorsiflexion and plantarflexion for females (left) and males (right). Each circle represents the MVT value for one participant, and horizontal black lines indicate the group mean. Circle size corresponds with Age, and circle color in the MS group corresponds to the participant EDSS score. We compared maximal strength between groups for dorsiflexion and plantarflexion separately using linear regression with a fixed effect of *Group* and covariates of *Age_c* (grand mean-centered *Age*) and *Sex*. Maximal strength was significantly lower for the MS group compared with the control group for both dorsiflexion (30.1 Nm vs. 36.7 Nm, *p* = 0.0008) and plantarflexion (86.2 Nm vs. 108.8 Nm, *p* = 0.0004). For the five participants whose TA data could not be included, their maximal strength values, reasons for exclusion, and sex were as follows: 23.9 Nm (insufficient yield, F), 38.5 Nm (insufficient yield, F), 0.75 Nm (weakness, M), 0.00 Nm (weakness, F), 0.78 Nm (weakness, F).

### 11.2. Group mean differences: linear mixed effect model structure and statistical output

**Figure A2.**
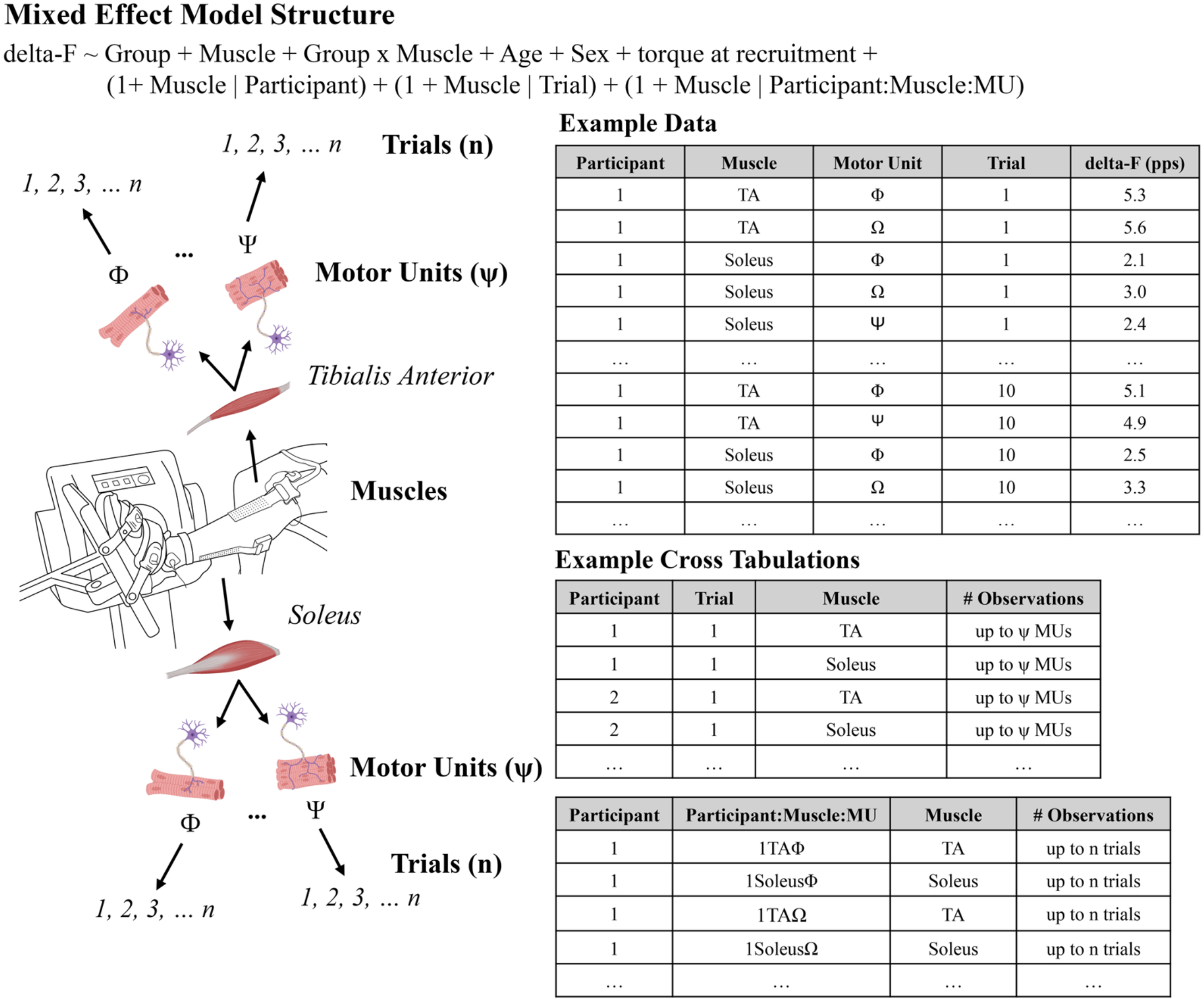
Schematic of the mixed-effect model structure for neurophysiological variables collected at the motor unit level. For any given outcome (e.g., delta F), each subject has multiple observations per motor unit and trial. Thus, there are three sources of variance we generally want to account for in the random effects (participant, trial, and motor unit). However, motor unit labels are arbitrary: i.e., motor unit “Φ” is same for one participant from trial to trial, but Φ in the soleus is *not* the same as Φ in the TA, and Φ for Participant 1 is not the same as Φ for Participant 2. Thus, we concatenate participant, muscle, and motor unit labels (e.g., “1TAΦ”), to obtain a random intercept that appropriately nests motor units within a given participant and muscle. Similar to the previous model, fixed effects then allow us to test the study’s main hypotheses with respect to Group and Group x Muscle. Continuous fixed effects were mean centered and categorical fixed effects were contrast coded using orthogonal polynomials. Notably, torque at recruitment was grand-mean centered (TQ_recrt_c), as opposed to being centered within a participant, and serves a motor unit-level covariate (except for the model with TQ_recrt as the outcome measure).

**Table A1.** Linear mixed effect model results for the composite variables and reverse engineering features: Effects of *Group, Muscle, Group* x *Muscle*

|  | Fixed effects of interest |  |  |
| --- | --- | --- | --- |
|  | *Group* | *Muscle* | *Group x Muscle* |
| Excitation composite | 0.11 [-0.02, 0.23]<br>F(1,102) = 2.7<br>*p* = 0.10 | -0.11 [-0.21, -0.005]<br>F(1,103) = 4.2<br>***p* = 0.04** | -0.22 [-0.42, -0.009]<br>F(1,103) = 4.2<br>***p* = 0.04** |
| Inhibition composite | -0.17 [-0.33, -0.005]<br>F(1,115) = 4.0<br>***p* = 0.048** | -0.05 [-0.18, 0.07]<br>F(1,115) = 0.74<br>*p* = 0.39 | -0.11 [-0.36, 0.14]<br>F(1,115) = 0.74<br>*p* = 0.39 |
| Neuromodulation composite | -0.05 [-0.17, 0.06]<br>F(1,119) = 0.78<br>*p* = 0.38 | -0.07 [-0.16, 0.01]<br>F(1,119) = 2.6<br>*p* = 0.11 | -0.14 [-0.32, 0.29]<br>F(1,119) = 2.6<br>*p* = 0.11 |
| Firing rates composite | -0.60 [-0.96, -0.24]<br>F(1,114) = 10.3<br>***p* = 0.002** | 0.04 [-0.17, 0.26]<br>F(1,114) = 0.16<br>*p* = 0.69 | 0.09 [-0.34, 0.52]<br>F(1,114) = 0.16<br>*p* = 0.69 |
| Braceheight | -0.007 [-0.03, 0.01]<br>*p* = 0.51 | -0.006 [-0.02, 0.01]<br>*p* = 0.50 | -0.01 [-0.05, 0.02]<br>*p* = 0.41 |
| Rate attenuation slope | -0.006 [-0.04, 0.02]<br>*p* = 0.70 | 0.10 [0.08, 0.12]<br>***p* < 0.0001** | 0.002 [-0.04, 0.05]<br>*p* = 0.93 |
| Delta-F | -0.51 [-0.90, -0.13]<br>***p* = 0.012** | 1.90 [1.67, 2.14]<br>***p* < 0.0001** | -0.49 [-0.95, -0.03]<br>***p* = 0.038** |
| Norm delta-F | -0.02 [-0.05, 0.01]<br>*p* = 0.17 | 0.11 [0.09, 0.13]<br>***p* < 0.0001** | -0.02 [-0.06, 0.01]<br>*p* = 0.23 |
| Recruitment torque | -0.24 [-0.93, 0.46]<br>*p* = 0.51 | 0.90 [0.21, 1.56]<br>***p* = 0.016** | 0.85 [-0.35, 2.05]<br>*p* = 0.17 |
| De-recruitment torque | -0.38 [-1.24, 0.48]<br>*p* = 0.39 | -2.97 [-3.67, -2.27]<br>***p* < 0.0001** | 1.02 [-0.35, 2.40]<br>*p* = 0.15 |
| Duration | 0.49 [-0.49, 1.47]<br>*p* = 0.33 | 2.60 [1.44, 3.78]<br>***p* = 0.0008** | -0.45 [-1.83, 0.92]<br>*p* = 0.51 |
| Recruitment Range | -1.30 [-2.17, -0.44]<br>***p* = 0.004** | -0.11 [-1.27, 1.06]<br>*p* = 0.84 | -0.50 [-1.90, 0.91]<br>*p* = 0.48 |
| Firing rate at recruitment | -0.55 [-0.91, -0.18]<br>***p* = 0.004** | 2.31 [2.08, 2.53]<br>***p* < 0.0001** | -0.29 [-0.74, 0.16]<br>*p* = 0.21 |
| Firing rate range | -0.93 [-1.42, -0.45]<br>***p* = 0.0003** | 3.57 [3.22, 3.92]<br>***p* < 0.0001** | -0.62 [-1.32, 0.08]<br>*p* = 0.09 |
| Maximal firing rate | -1.16 [-1.80, -0.53]<br>***p* = 0.001** | 4.97 [4.55, 5.40]<br>***p* < 0.0001** | -0.61 [-1.46, 0.25]<br>*p* = 0.18 |
| Firing rate at de-recruitment | -0.09 [-0.36, 0.18]<br>$p = 0.52$ | 1.64 [1.45, 1.83]<br>$p < \mathbf{0.0001}$ | 0.09 [-0.29, 0.47]<br>$p = 0.65$ |

**Table A2.**
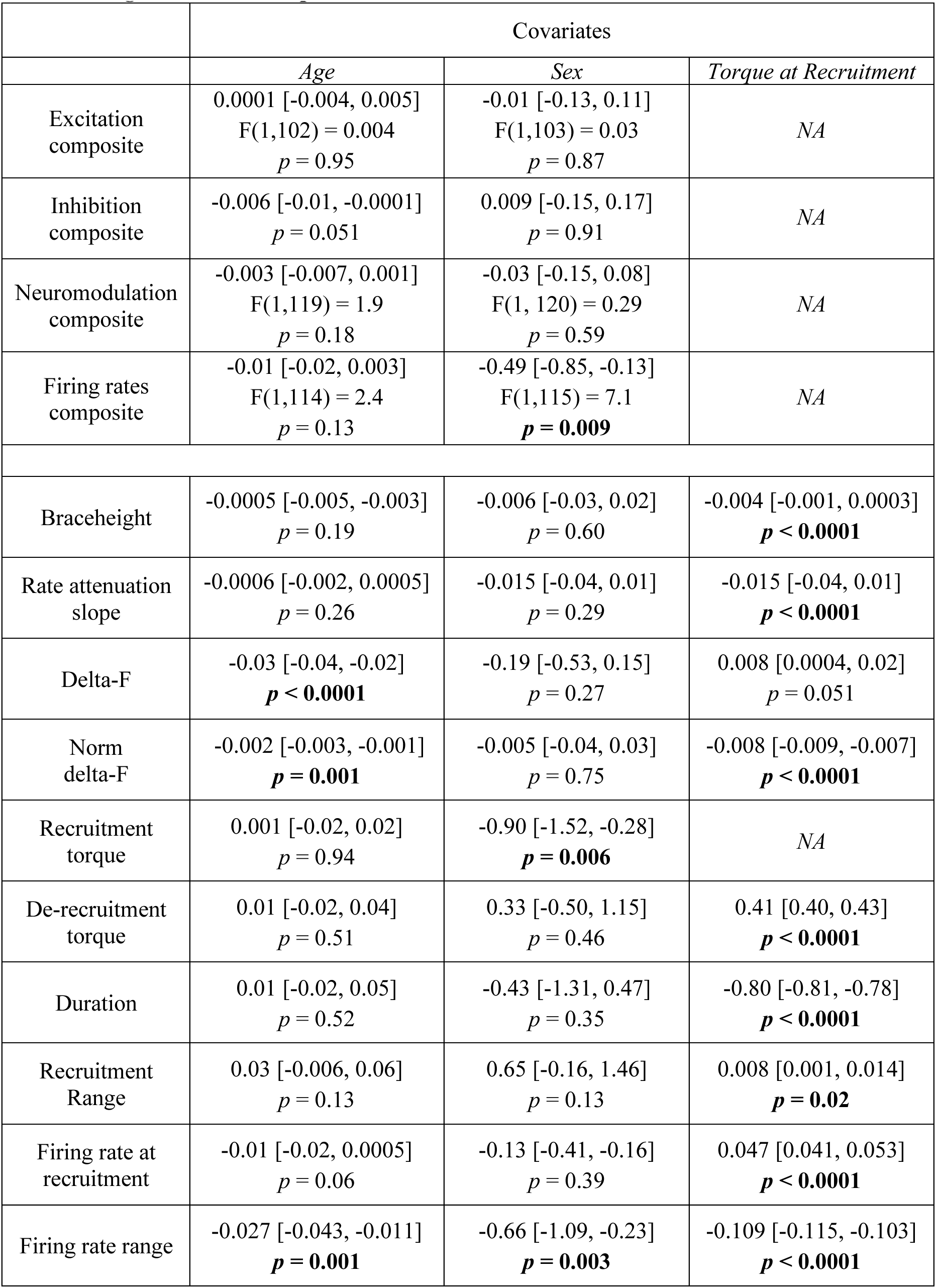
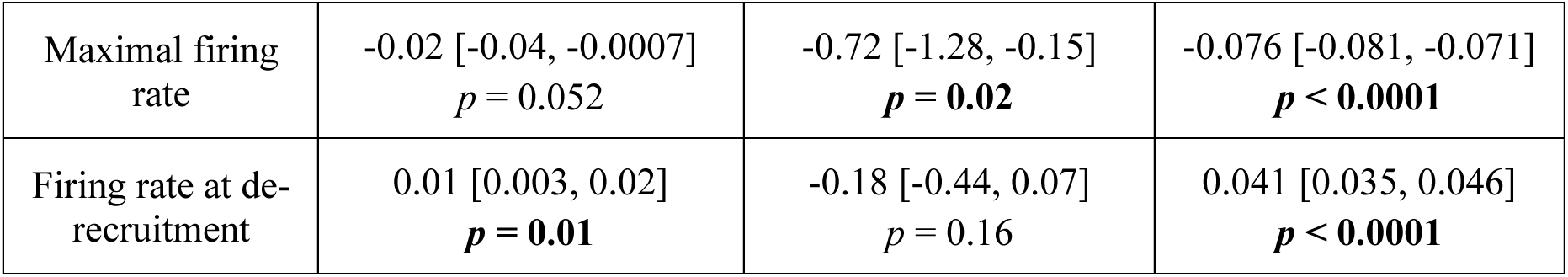
Linear mixed effect model results for the composite variables and reverse engineering features: Effects of *Age, Sex,* and *Torque at Recruitment*

|  | Covariates |  |  |
| --- | --- | --- | --- |
|  | *Age* | *Sex* | *Torque at Recruitment* |
| Excitation composite | 0.0001 [-0.004, 0.005]<br>F(1,102) = 0.004<br>$p = 0.95$ | -0.01 [-0.13, 0.11]<br>F(1,103) = 0.03<br>$p = 0.87$ | NA |
| Inhibition composite | -0.006 [-0.01, -0.0001]<br>$p = 0.051$ | 0.009 [-0.15, 0.17]<br>$p = 0.91$ | NA |
| Neuromodulation composite | -0.003 [-0.007, 0.001]<br>F(1,119) = 1.9<br>$p = 0.18$ | -0.03 [-0.15, 0.08]<br>F(1, 120) = 0.29<br>$p = 0.59$ | NA |
| Firing rates composite | -0.01 [-0.02, 0.003]<br>F(1,114) = 2.4<br>$p = 0.13$ | -0.49 [-0.85, -0.13]<br>F(1,115) = 7.1<br>$p = \mathbf{0.009}$ | NA |
| Braceheight | -0.0005 [-0.005, -0.003]<br>$p = 0.19$ | -0.006 [-0.03, 0.02]<br>$p = 0.60$ | -0.004 [-0.001, 0.0003]<br>$p < \mathbf{0.0001}$ |
| Rate attenuation slope | -0.0006 [-0.002, 0.0005]<br>$p = 0.26$ | -0.015 [-0.04, 0.01]<br>$p = 0.29$ | -0.015 [-0.04, 0.01]<br>$p < \mathbf{0.0001}$ |
| Delta-F | -0.03 [-0.04, -0.02]<br>$p < \mathbf{0.0001}$ | -0.19 [-0.53, 0.15]<br>$p = 0.27$ | 0.008 [0.0004, 0.02]<br>$p = 0.051$ |
| Norm delta-F | -0.002 [-0.003, -0.001]<br>$p = \mathbf{0.001}$ | -0.005 [-0.04, 0.03]<br>$p = 0.75$ | -0.008 [-0.009, -0.007]<br>$p < \mathbf{0.0001}$ |
| Recruitment torque | 0.001 [-0.02, 0.02]<br>$p = 0.94$ | -0.90 [-1.52, -0.28]<br>$p = \mathbf{0.006}$ | NA |
| De-recruitment torque | 0.01 [-0.02, 0.04]<br>$p = 0.51$ | 0.33 [-0.50, 1.15]<br>$p = 0.46$ | 0.41 [0.40, 0.43]<br>$p < \mathbf{0.0001}$ |
| Duration | 0.01 [-0.02, 0.05]<br>$p = 0.52$ | -0.43 [-1.31, 0.47]<br>$p = 0.35$ | -0.80 [-0.81, -0.78]<br>$p < \mathbf{0.0001}$ |
| Recruitment Range | 0.03 [-0.006, 0.06]<br>$p = 0.13$ | 0.65 [-0.16, 1.46]<br>$p = 0.13$ | 0.008 [0.001, 0.014]<br>$p = \mathbf{0.02}$ |
| Firing rate at recruitment | -0.01 [-0.02, 0.0005]<br>$p = 0.06$ | -0.13 [-0.41, -0.16]<br>$p = 0.39$ | 0.047 [0.041, 0.053]<br>$p < \mathbf{0.0001}$ |
| Firing rate range | -0.027 [-0.043, -0.011]<br>$p = \mathbf{0.001}$ | -0.66 [-1.09, -0.23]<br>$p = \mathbf{0.003}$ | -0.109 [-0.115, -0.103]<br>$p < \mathbf{0.0001}$ |
| Maximal firing rate | -0.02 [-0.04, -0.0007]<br>$p = 0.052$ | -0.72 [-1.28, -0.15]<br>$p = \mathbf{0.02}$ | -0.076 [-0.081, -0.071]<br>$p < \mathbf{0.0001}$ |
| Firing rate at de-recruitment | 0.01 [0.003, 0.02]<br>$p = \mathbf{0.01}$ | -0.18 [-0.44, 0.07]<br>$p = 0.16$ | 0.041 [0.035, 0.046]<br>$p < \mathbf{0.0001}$ |

**Table A3.**
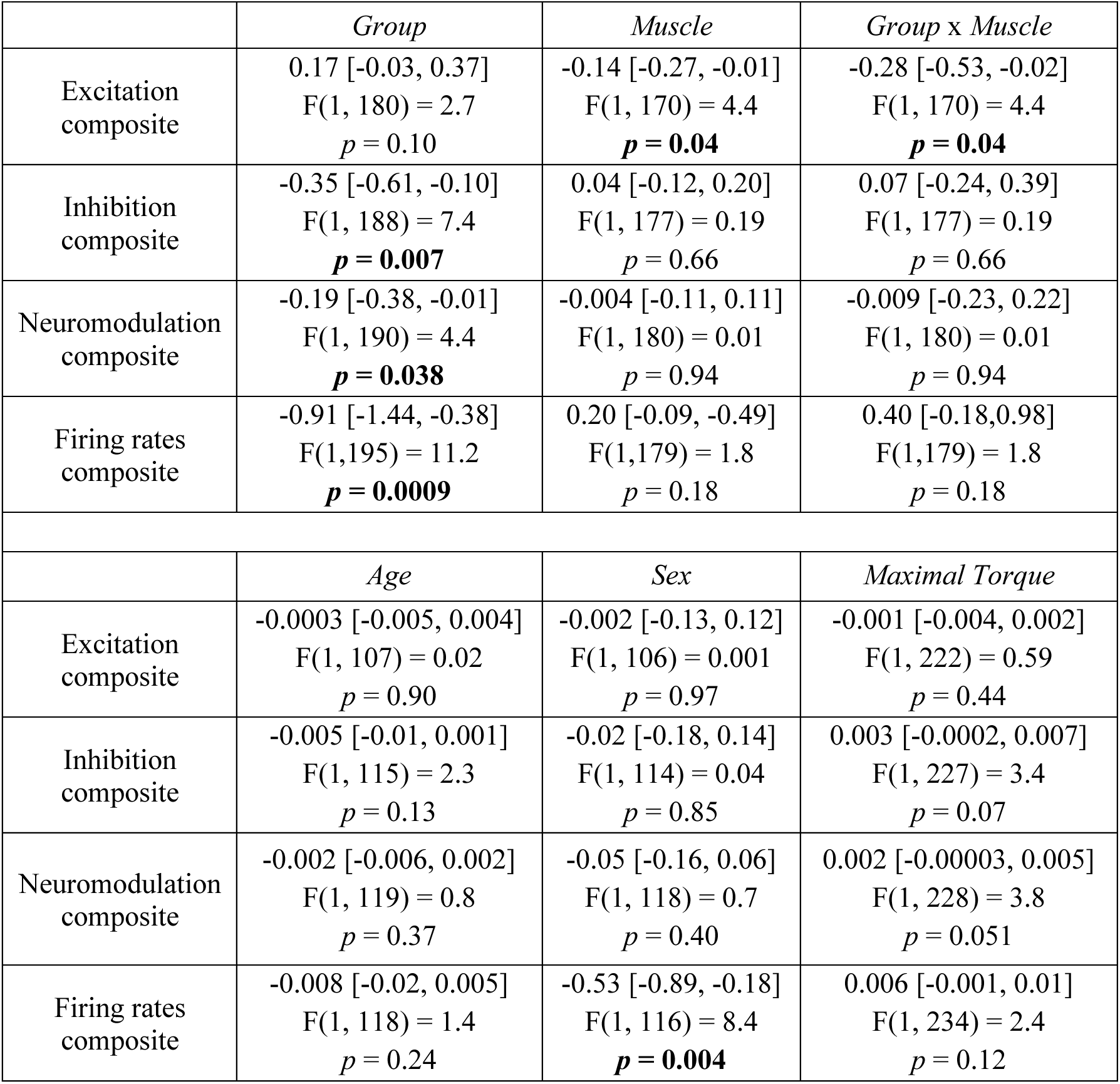
Linear mixed effect model results for the composite variables when controlling for maximal strength.

|  | *Group* | *Muscle* | *Group x Muscle* |
| --- | --- | --- | --- |
| Excitation composite | 0.17 [-0.03, 0.37]<br>F(1, 180) = 2.7<br>$p = 0.10$ | -0.14 [-0.27, -0.01]<br>F(1, 170) = 4.4<br>$p = \mathbf{0.04}$ | -0.28 [-0.53, -0.02]<br>F(1, 170) = 4.4<br>$p = \mathbf{0.04}$ |
| Inhibition composite | -0.35 [-0.61, -0.10]<br>F(1, 188) = 7.4<br>$p = \mathbf{0.007}$ | 0.04 [-0.12, 0.20]<br>F(1, 177) = 0.19<br>$p = 0.66$ | 0.07 [-0.24, 0.39]<br>F(1, 177) = 0.19<br>$p = 0.66$ |
| Neuromodulation composite | -0.19 [-0.38, -0.01]<br>F(1, 190) = 4.4<br>$p = \mathbf{0.038}$ | -0.004 [-0.11, 0.11]<br>F(1, 180) = 0.01<br>$p = 0.94$ | -0.009 [-0.23, 0.22]<br>F(1, 180) = 0.01<br>$p = 0.94$ |
| Firing rates composite | -0.91 [-1.44, -0.38]<br>F(1,195) = 11.2<br>$p = \mathbf{0.0009}$ | 0.20 [-0.09, -0.49]<br>F(1,179) = 1.8<br>$p = 0.18$ | 0.40 [-0.18,0.98]<br>F(1,179) = 1.8<br>$p = 0.18$ |
|  | *Age* | *Sex* | *Maximal Torque* |
| Excitation composite | -0.0003 [-0.005, 0.004]<br>F(1, 107) = 0.02<br>$p = 0.90$ | -0.002 [-0.13, 0.12]<br>F(1, 106) = 0.001<br>$p = 0.97$ | -0.001 [-0.004, 0.002]<br>F(1, 222) = 0.59<br>$p = 0.44$ |
| Inhibition composite | -0.005 [-0.01, 0.001]<br>F(1, 115) = 2.3<br>$p = 0.13$ | -0.02 [-0.18, 0.14]<br>F(1, 114) = 0.04<br>$p = 0.85$ | 0.003 [-0.0002, 0.007]<br>F(1, 227) = 3.4<br>$p = 0.07$ |
| Neuromodulation composite | -0.002 [-0.006, 0.002]<br>F(1, 119) = 0.8<br>$p = 0.37$ | -0.05 [-0.16, 0.06]<br>F(1, 118) = 0.7<br>$p = 0.40$ | 0.002 [-0.00003, 0.005]<br>F(1, 228) = 3.8<br>$p = 0.051$ |
| Firing rates composite | -0.008 [-0.02, 0.005]<br>F(1, 118) = 1.4<br>$p = 0.24$ | -0.53 [-0.89, -0.18]<br>F(1, 116) = 8.4<br>$p = \mathbf{0.004}$ | 0.006 [-0.001, 0.01]<br>F(1, 234) = 2.4<br>$p = 0.12$ |

### 11.3. Group differences in distribution spread and shape: statistical output

**Table A4.** Tests for equality of variance and differences in distribution shape for the composite variables.

|  | TA |  | Soleus |  |
| --- | --- | --- | --- | --- |
|  | Levene's test | *Komolgorov-Smirnov test* | Levene's test | *Komolgorov-Smirnov test* |
| Excitation | 0.17 vs. 0.09<br>$p = \mathbf{0.048}$<br>F(1,115) = 4.0 | D = 0.41<br>$p < \mathbf{0.0001}$ | 0.21 vs. 0.08<br>$p = \mathbf{0.019}$<br>F(1,121) = 5.6 | D = 0.53<br>$p < \mathbf{0.0001}$ |
| Inhibition | 0.35 vs. 0.10<br>**$p = 0.002$**<br>$F(1,115) = 10.1$ | $D = 0.55$<br>**$p < 0.0001$** | 0.27 vs. 0.19<br>$p = 0.68$<br>$F(1,121) = 0.16$ | $D = 0.41$<br>**$p = 0.0001$** |
| Neuromodulation | 0.19 vs. 0.06<br>**$p = 0.004$**<br>$F(1,115) = 8.43$ | $D = 0.54$<br>**$p < 0.0001$** | 0.13 vs. 0.07<br>$p = 0.30$<br>$F(1,121) = 1.1$ | $D = 0.51$<br>**$p < 0.0001$** |
| Firing rates | 1.07 vs. 0.72<br>$p = 0.51$<br>$F(1,116) = 0.44$ | $D = 0.29$<br>**$p = 0.008$** | 1.59 vs. 0.62<br>**$p = 0.044$**<br>$F(1,121) = 4.1$ | $D = 0.63$<br>**$p < 0.0001$** |

**Table A5.** Tests for equality of variance for the reverse engineering features.

|  | TA | Soleus |
| --- | --- | --- |
| Braceheight | 0.009 vs. 0.004<br>**$p = 0.037$**<br>$F(1,116) = 4.4$ | 0.005 vs. 0.002<br>$p = 0.051$<br>$F(1,121) = 3.9$ |
| Rate attenuation slope | 0.02 vs. 0.01<br>$p = 0.10$<br>$F(1,116) = 6.4$ | 0.006 vs. 0.004<br>$p = 0.52$<br>$F(1,121) = 0.4$ |
| delta-F | 2.4 vs. 1.2<br>**$p = 0.013$**<br>$F(1,115) = 6.4$ | 1.17 vs. 0.90<br>$p = 0.22$<br>$F(1,121) = 1.5$ |
| Normalized delta-F | 0.02 vs. 0.007<br>$p = 0.076$<br>$F(1,115) = 3.2$ | 0.02 vs. 0.01<br>$p = 0.18$<br>$F(1,121) = 1.8$ |
| Recruitment torque | 11.7 vs. 7.5<br>$p = 0.19$<br>$F(1,116) = 1.8$ | 6.4 vs. 5.0<br>$p = 0.29$<br>$F(1,121) = 1.1$ |
| De-recruitment torque | 9.9 vs. 4.1<br>**$p = 0.01$**<br>$F(1,116) = 6.9$ | 13.4 vs. 6.3<br>**$p = 0.03$**<br>$F(1,121) = 4.9$ |
| Duration | 13.2 vs. 3.1<br>**$p = 0.007$**<br>$F(1,116) = 7.7$ | 12.1 vs. 3.2<br>**$p = 0.049$**<br>$F(1,121) = 4.0$ |
| Recruitment Range | 13.0 vs. 6.6<br>**$p = 0.009$**<br>$F(1,116) = 7.0$ | 7.1 vs. 3.7<br>**$p = 0.048$**<br>$F(1,121) = 4.0$ |
| Firing rate at recruitment | 2.0 vs. 1.1<br>$p = 0.35$<br>$F(1,116) = 0.9$ | 0.6 vs. 0.3<br>$p = 0.11$<br>$F(1,121) = 2.5$ |
| Firing rate range | 4.1 vs 3.0<br>$p = 0.46$<br>$F(1,116) = 0.6$ | 1.3 vs. 0.8<br>$p = 0.12$<br>$F(1,121) = 2.4$ |
| Maximal firing rate | 6.4 vs. 3.8<br>$p = 0.37$<br>$F(1,116) = 0.8$ | 2.3 vs. 1.2<br>**$p = 0.049$**<br>$F(1,121) = 4.0$ |
| Firing rate at de-recruitment | 1.15 vs. 0.97<br>$p = 0.57$<br>$F(1,116) = 0.3$ | 0.8 vs. 0.2<br>$p = 0.16$<br>$F(1,121) = 2.0$ |

